# Investigating the Interplay Between Prematurity and Genetic Variation in the Context of Rare Developmental Disorders

**DOI:** 10.1101/2025.03.19.25324255

**Authors:** Olivia Wootton, Patrick Campbell, Sarah Richardson, Sarah J Lindsay, Qin Qin Huang, Erwan Delage, Sana Amanat, Hilary S Wong, Matthew E Hurles, Michael A Simpson, Elizabeth J Radford, Hilary C Martin

## Abstract

Rare damaging genetic variation accounts for a substantial proportion of the risk of rare developmental disorders (DDs), but common genetic variants as well as environmental factors, including prematurity, also contribute. Little is known about the interplay between prematurity and genetic variation in influencing phenotypic outcomes in DDs, nor about how genetic factors may contribute to risk of preterm birth in DDs. To address these questions, we leverage phenotypic and genetic data from 21,712 patients with DDs recruited for clinical sequencing, 16% of whom were born prematurely. We find that prematurity is associated with more severe clinical phenotypes amongst these DD patients, including more affected organ systems and more delayed developmental milestones, with prematurity and monogenic diagnoses contributing independently and additively to phenotypic severity. We identify genes and gene sets enriched for diagnostic mutations amongst preterm children with DDs. We also demonstrate an enrichment of *de novo* mutations (DNMs) in both term and preterm probands; the fraction of cases explained by DNMs in known DD-associated genes is higher in term than preterm cases (25% versus 20%) but DNMs in as-yet-undiscovered genes likely contribute approximately equally to both groups (14% versus 13%). Finally, we show that the positive association between polygenic predisposition to education-related traits and gestational duration is likely to be the result of genetically-influenced parental traits or confounders, rather than direct genetic effects in the child, and that the presence of a monogenic diagnosis modifies this association. Our findings emphasise the importance of considering environmental exposures like prematurity in understanding outcomes in DDs suspected to have a genetic component, and motivate further exploration of the role that genetic variation plays in influencing prematurity.

## Introduction

Understanding how the interplay of genetics and environment influences phenotype is an enduring question in biology. In the context of rare developmental disorders (DDs), there is evidence that both rare^1^ and common^2,3^ genetic variation as well as environmental factors contribute to aetiology.^4^ DDs are a group of conditions that arise in embryonic life or during early development and alter the developmental trajectory. Research over the past decade has had considerable success in identifying the rare genetic variation that causes many of these conditions. At present, a diagnostic rare variant is found in 25-45% of individuals with DDs who are referred for exome or genome sequencing.^5–7^ One environmental factor likely to also contribute to phenotypes among these individuals is prematurity, given premature delivery occurs in 9.9% of births^8^ and a common sequelae is developmental delay.^9^ This hypothesis is supported by recent work from the Deciphering Developmental Disorders (DDD) study, a large cohort of children with DDs, which demonstrated that preterm infants (who were enriched in the cohort, comprising 16% of probands) were less likely to receive a monogenic diagnosis than term infants.^5^ To date, however, there has been limited investigation into the role of prematurity among children with DDs with a suspected genetic component.

The impact of prematurity on phenotypic presentation of DDs is important from a clinical perspective. It is well established that individuals with pathogenic variants in more than one Mendelian-acting gene can have composite phenotypes.^10–12^ Similarly, an individual with a pathogenic variant who is also born prematurely may have a composite phenotype. Consistent with this, recent research has found that in autism, a common neurodevelopmental condition with genetic contributions, individuals diagnosed with the condition have more comorbidities if born prematurely than if born at term.^13^ In rare DDs, teasing apart the phenotypic impact of prematurity from that of underlying monogenic diagnoses could provide clinicians and families with a more complete explanation of an individual’s clinical features.

Although preterm birth is generally considered to be an ‘environmental’ perturbation, a growing body of evidence shows that it is influenced by genetic variation, both maternal and fetal.^14,15^ Studies have found higher rates of *de novo* variants in preterm infants, linking affected genes to fetal brain development^16^, and have implicated structural variants that are known to cause developmental delay^17^. Additionally, the clinical synopsis of several DDs in Online Mendelian Inheritance in Man include premature birth as a feature (e.g. MIM: 619488, 618737, 620155). However, given the small number of cases these data are drawn from,^18–20^ the extent to which rare pathogenic variants in these genes are robustly associated with an increased risk of prematurity is unknown. Furthermore, genetic variation that contributes to prematurity may affect developmental trajectories in children with DDs. Recent work has found that common genetic variant predisposition to preterm birth is negatively genetically correlated with years in education (educational attainment; EA) and positively genetically correlated with risk of neurodevelopmental conditions.^3^ Consistent with these findings, preterm probands in the DDD study were found to have a polygenic predisposition to lower EA compared to term probands.^3^ It is unclear whether polygenic predisposition for lower EA has a direct genetic effect on an individual’s risk of being born prematurely (i.e. the effects of genetic variants in an individual on that individual’s own phenotype), as opposed to an indirect genetic effect that may be mediated through parental behaviours or the prenatal environment,. Regardless, previous studies suggest that genetic risk for prematurity and developmental disorders may overlap, and that common and rare genetic variation associated with prematurity may confound associations between prematurity and phenotypic outcomes in DDs.

Here, we use data from large cohorts of individuals with developmental disorders recruited for clinical sequencing (N=21,712) to investigate three questions. First, we assess the phenotypic impact of prematurity on clinical features and developmental outcomes. Second, we test whether there are DD-associated genes and disease-associated gene sets in which diagnostic variants are enriched among individuals with DDs born prematurely, and we evaluate the overall contribution of *de novo* variation to DDs in term and preterm probands. Third, we explore whether established associations between common variant predisposition to education-related traits and gestational duration are modified by the presence of a monogenic diagnosis and whether these associations are mediated through direct genetic effects. In addressing these questions, this work aims to improve our scientific and clinical understanding of the interplay between genetic variation and prematurity on rare DDs.

## Results

Our analysis included probands with data on gestational age at birth from the DDD study (N=13,401) as well as DDD-like probands from the rare disease programme of the 100,000 Genomes Project (100kGP; N=8,311). Gestational age groups were classified according to the World Health Organization definitions of prematurity: extreme (<28 weeks), very (28 to <32 weeks), and moderate (32 to <37 weeks).^8,21^ Probands were further categorised as “diagnosed” or “undiagnosed”. In DDD, “diagnosed” probands had a clinically confirmed genetic diagnosis or predicted pathogenic or likely pathogenic variant. In 100kGP, “diagnosed” probands included those with a clinically confirmed diagnostic variant or prioritized variants thought highly likely to be diagnostic that were awaiting clinical review (See <u>Methods</u> for further details). **Table 1** provides an overview of the basic demographic and clinical characteristics of probands from each cohort.

**Table 1.** Descriptive statistics of the datasets combined.

|  | **DDD (N=13,401)** | **100kGP (N=8,311)** |
| --- | --- | --- |
| **Gestational Age Group (%)** |  |  |
| Term<br>(37+ weeks) | 11,246 (83.9%) | 7,042 (84.7%) |
| Moderately Premature<br>(32-36 weeks) | 1,753 (13.1%) | 998 (12%) |
| Very Premature<br>(28-31 weeks) | 291 (2.2%) | 205 (2.5%) |
| Extremely Premature<br>(<28 weeks) | 111 (0.8%) | 66 (0.8%) |
| **Number diagnosed (%)** | 5,493 (41%) | 2383 (28.7%) |
| Term | 4750 (42.2%) | 2092 (29.7%) |
| Moderately Premature | 627 (35.8%) | 243 (24.3%) |
| Very Premature | 91 (31.3%) | 41 (20%) |
| Extremely Premature | 25 (22.5%) | 7 (10.6%) |
| **Age at recruitment in years**<br>**(mean ± SD)** | 7.32 ± 6.13 | 7.87 ± 5.14 |
| **Number in trios (%)** | 9,829 (73.4%) | 5,585 (67.2%) |
| **Birthweight in grams (mean ± SD)** | 3,047 ± 772 | 3,114 ± 732 |
| **Number of organ systems affected<sup>†</sup>** | 4.25 ± 2.15 | 3.52 ± 2.34 |

<sup>†</sup> As terms may be associated with multiple organ systems, the number of distinct affected organ systems was calculated using the approach outlined in Niemi *et al.* to assign each HPO term to just one organ system.<sup>2</sup>

### Premature birth affects clinical features among individuals with developmental disorders suspected to have a genetic component

We first sought to test whether premature birth influences phenotypic presentation in children with DDs and whether additional genetic factors, such as diagnostic variants and polygenic predisposition, also contribute. To investigate these relationships in DDs, we tested for associations between the degree of prematurity and phenotypic outcomes derived from clinical records, HPO terms, and developmental milestone data. We further tested whether these associations were modified by the presence of a monogenic diagnosis. The results for the association analysis were generally consistent for probands from DDD and 100kGP, thus results from the meta-analysis are presented below (See **Supplementary Figures 1-3** for cohort-specific results).

We started by estimating the association between degree of prematurity and selected antenatal factors, recapitulating established associations observed in the general population.^22–25^ Specifically, we found that premature delivery was more likely for mothers with a history of previous pregnancy loss, diabetes during pregnancy, an abnormal antenatal ultrasound scan and in multifetal pregnancies (**Supplementary Figure 1**). Multifetal pregnancy showed a strong, significant association with all categories of prematurity with the greatest odds observed for extremely preterm delivery (OR=14.03, p=7.32×10^-36^). Maternal diabetes was associated with a significantly increased likelihood of moderate prematurity (OR=2.10, p=1.78×10^-16^), but not very or extreme prematurity. We found that a genetic diagnosis does not change the relationship between prematurity and these antenatal factors (**Supplementary Table 2**).

Next, we assessed associations between preterm birth and phenotypes recorded for probands from both cohorts, including HPO terms describing affected organs and/or organ systems and the severity of intellectual and/or developmental delay. On average, premature probands across all categories of prematurity were more likely to have lower birth weight (p<3.23×10^-7^) and had more affected organ systems than term probands (p<1.91×10^-6^) (**Supplementary Figure 2**). Reassuringly, we found significantly increased odds of being assigned the HPO term, “abnormality of prenatal development or birth”, for all categories of prematurity, with the odds increasing as the degree of prematurity increased (OR=3.48-10.79, p<2.75×10^-45^). All categories of premature birth were significantly associated with increased odds of being assigned an HPO term related to abnormality of the cardiovascular and digestive systems (p < 6.90×10^-5^)(**Figure 1**; **Supplementary Figure 3**). For these organ systems, very preterm children had higher odds than moderately preterm children. However, we did not observe increased odds for extremely preterm children relative to other categories of prematurity. The confidence intervals for the extremely preterm estimates were wide, likely due to the small number of extremely preterm probands in the analysis. We found no significant associations between any category of prematurity and severity of intellectual disability or developmental delay or abnormalities of the ears, eyes, immune system, integument, limbs, and nervous system (**Supplementary** Figures 2-3).

**Figure 1.**
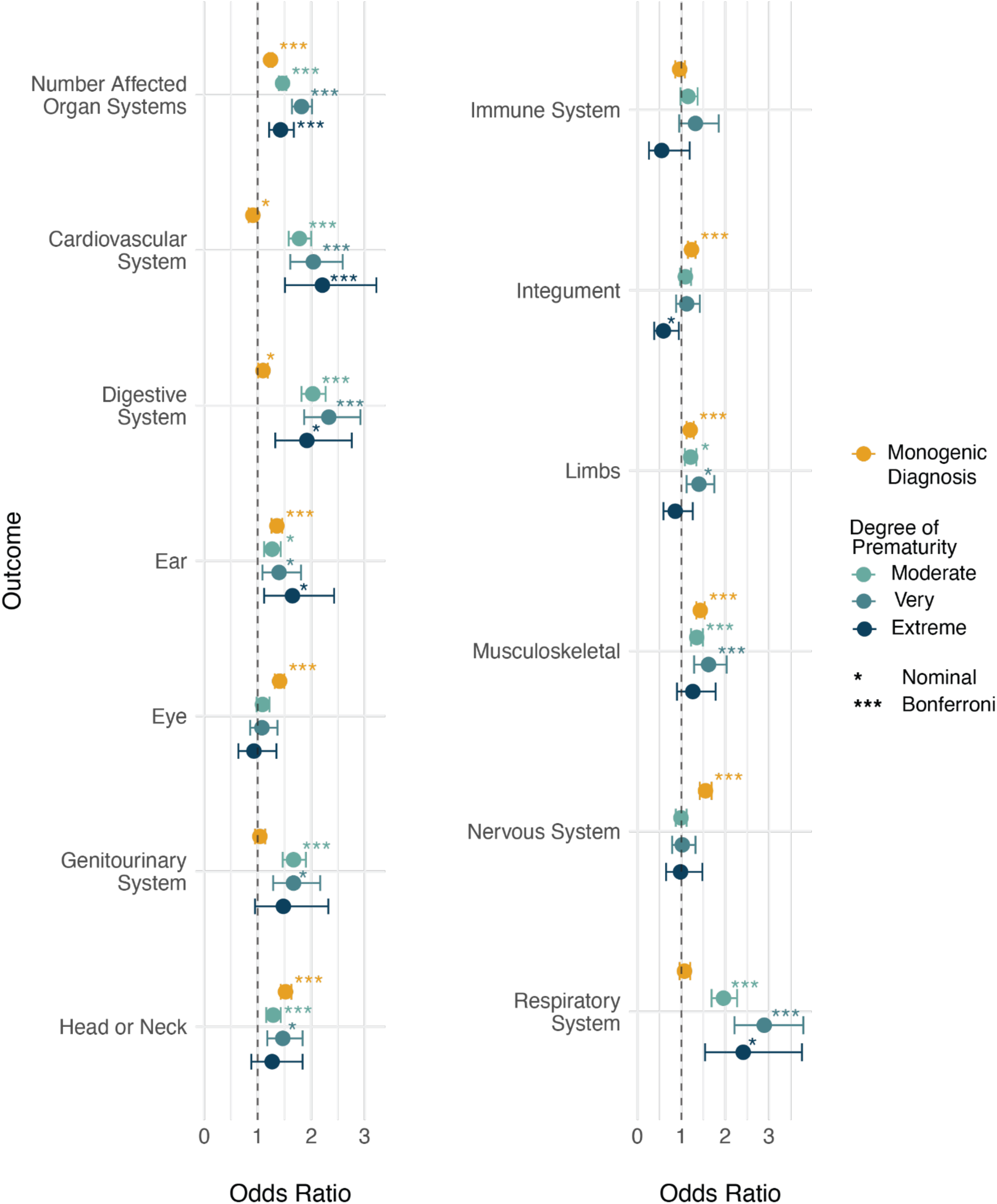
Associations between phenotypic abnormalities, degree of prematurity, and the presence of a monogenic diagnosis. The figure illustrates the odds of being assigned a term describing a phenotypic abnormality of an organ or organ system, or a descendant HPO term, for probands in each category of prematurity compared to those born at term (blue circles) and for probands with a monogenic diagnosis compared to those without (yellow circles). Odds ratios were estimated from a model regressing phenotypic outcome on degree of prematurity and monogenic diagnosis. Error bars indicate 95% confidence intervals.

In an extended model that aimed to test if the presence of a monogenic diagnosis modified phenotypic associations with prematurity, we found that the organ systems impacted by prematurity differed from those associated with diagnostic variants (**Figure 1**). For instance, while the presence of a diagnostic variant did not increase the likelihood of abnormalities in the cardiovascular or digestive systems, it significantly increased the odds of abnormalities in the nervous system, eyes, ears, and integument (**Figure 1**). No significant interactions were detected between any category of prematurity and the presence of a monogenic diagnosis, suggesting that preterm birth and diagnostic variants independently and additively influence clinical features in developmental disorders (**Supplementary Table 3**).

We then considered two additional sets of HPO terms for this analysis: a set of prematurity-associated HPO terms chosen by clinicians (e.g., retinopathy of prematurity, intraventricular haemorrhage and necrotising enterocolitis) and a curated set of medically-relevant HPO terms selected for their relevance to neurodevelopmental disorders^2^. We found that few to no probands in these cohorts were assigned the prematurity-associated HPO terms, thus we did not conduct the association analysis (**Supplementary Figure 4**). When analysing the medically-relevant HPO terms, we saw few significant associations between prematurity and the phenotypes (**Supplementary Figure 5**). Two exceptions were that the likelihood of short stature was significantly higher for children born across all categories of prematurity compared to those born at term (p < 8.98×^-7^), and that moderate prematurity was associated with significantly reduced odds of being assigned an HPO term for seizures (OR=0.75, p=5.38×10^-7^) (**Supplementary Figure 5**). There were no significant interactions between degree of prematurity and the presence of a monogenic diagnosis for any of the medically-relevant HPO terms (**Supplementary Table 3**).

### Attainment of developmental milestones

In the analysis described above, we did not detect an association between degree of prematurity and severity of developmental delay or intellectual disability. However, this analysis relied on clinical reports of severity, which were missing for 75% of probands. To address this limitation, we utilised developmental milestone data, available in DDD probands, to reexamine the impact of prematurity and genetic variation on neurodevelopment. We first compared time taken to achieve a developmental milestone (social smile, sitting independently, walking independently, and first words) across gestational age groups using Kaplan-Meier survival analysis. Gestational age group was significantly associated with variation in the time taken to achieve all assessed developmental milestones (log rank test p < 2×10^-3^) (**Supplementary** Figure 6; **Supplementary Table 4**). Extremely preterm probands consistently achieved all milestones later than other groups. Term probands achieved the developmental milestones of “social smile” and “sitting independently” significantly earlier than probands from the preterm gestational age groups (**Supplementary Table 4**). A similar trend was seen for age of walking and age of first words, but only extremely preterm probands showed a significant delay compared to term probands (p < 1.73×10^-4^). We extended this analysis with Cox regression models to explore how the presence of a diagnostic variant influenced the time taken to achieve these developmental milestones. In these models, the presence of a genetic diagnosis was associated with increased time to milestone attainment across the gestational age groups for all developmental milestones (p < 0.04) (**Figure 2**). However, there was no evidence of an interaction between a monogenic diagnosis and gestational age at birth on the time to milestone attainment (**Supplementary Table 5**). These findings are consistent with the findings of our phenotype association analysis whereby prematurity and the diagnostic variant act additively as opposed to multiplicatively to influence outcomes in DDs.

**Figure 2.**
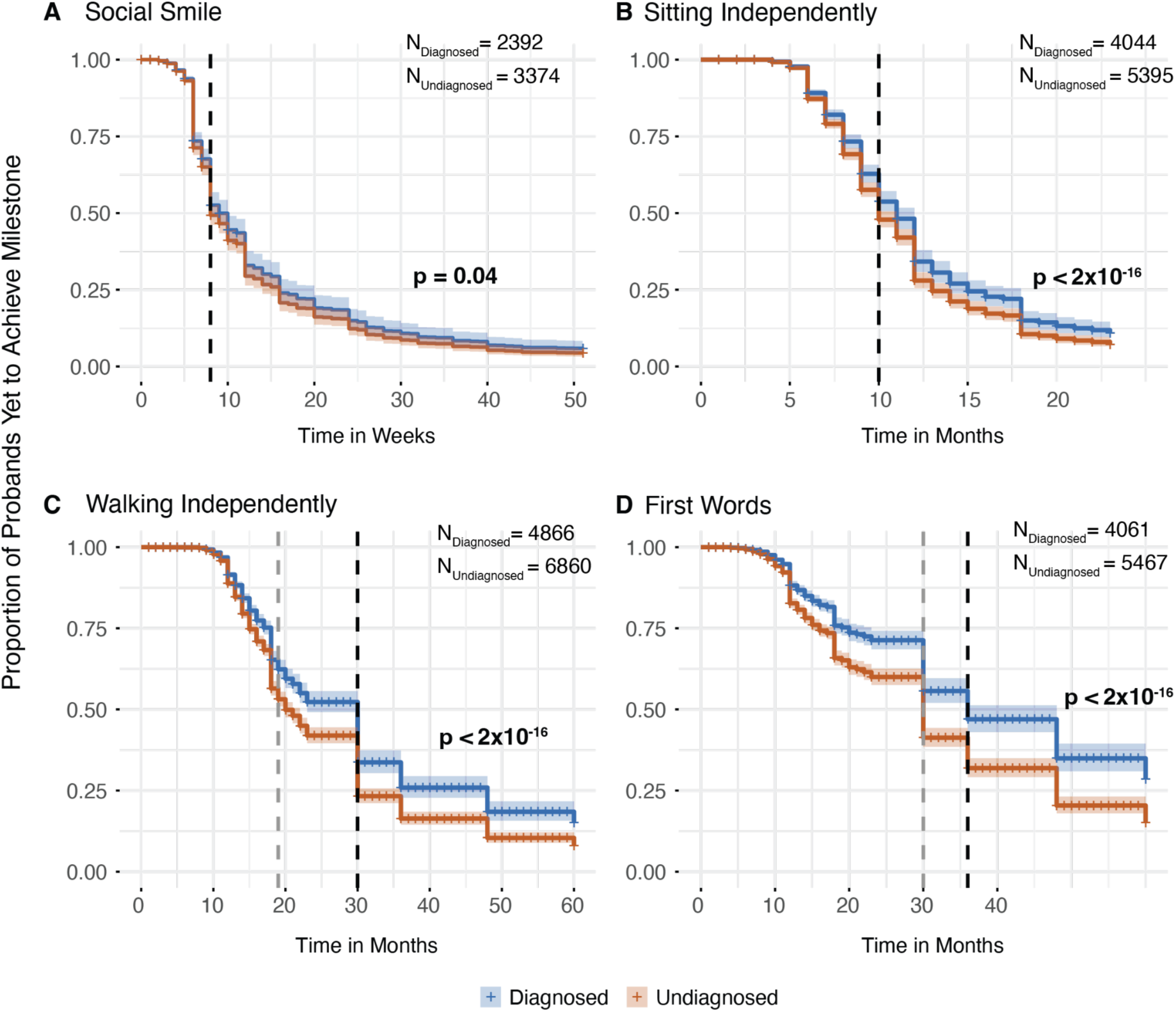
Survival curves comparing time taken to achieve developmental milestones for preterm probands with and without a genetic diagnosis. X-axis denotes time in weeks or months, 0 corresponds to date of birth, and the y-axis shows the proportion of probands who have not achieved the developmental milestone after a given time period. Dashed vertical line shows median time to achieve milestone for diagnosed term probands (black) and undiagnosed term probands (grey). (Note that this was the same for diagnosed and undiagnosed probands in panel A and B). Wald test p-values are for the effect of the presence of a monogenic diagnosis on time taken to achieve a developmental milestone if the gestational age group was held constant. **A)** Time (in weeks) taken to achieve the developmental milestone, social smile. **B**) Time (in months) taken to achieve the developmental milestone, sitting independently. **C**) Time (in months) taken to achieve the developmental milestone, walking independently. **D**) Time (in months) taken to achieve the developmental milestone, first words. Colour represents diagnostic status, shaded ribbons depict 95% confidence intervals.

### Differential enrichment of monogenic diagnoses in specific genes and gene sets in premature probands

As genetic diagnoses of DDs are increasingly being made *in utero* through prenatal sequencing,^26,27^ the identification of specific DD genes associated with prematurity could be valuable to refine preterm birth risk stratification and implement appropriate antenatal care in affected fetuses. We therefore investigated if diagnostic variants in any specific genes or gene sets were associated with prematurity in an analysis of 7,581 diagnosed probands (986 preterm) across both DDD and 100kGP.

For the gene-based analysis, we performed logistic regression to test if preterm birth was more likely for carriers of a diagnostic variant in one of the 44 genes selected for this analysis on the basis of having sufficient power (See <u>Supplementary Methods</u>). We identified four genes, *EP300* (OR=2.7, p=2.34×10^-3^), *KMT2A* (OR=1.9, p=5.54×10^-3^), *NSD1* (OR=2.2, p=4.96×10^-2^), and *NF1* (OR=2.4, p=1.81×10^-2^), in which a diagnostic mutation was associated with an increased likelihood of prematurity at nominal significance, and one gene, *MECP2* (OR=0.1, p=2.24×10^-2^), in which the likelihood was reduced (**Supplementary Table 6**). After correction for multiple testing, no associations remained significant. After limiting the analysis to probandswith diagnostic variants that had been clinically annotated as pathogenic or likely pathogenic (6,223 probands, 791 premature), we found that all identified genes except *NF1* remained nominally significantly associated with prematurity and that the association with *EP300* passed multiple testing correction (**Supplementary Table 7**).

Next, we conducted Fisher’s exact tests to assess differences in the proportion of diagnosed probands with a diagnostic mutation in selected gene sets across the categories of gestational duration. The analysis focused on four gene sets prioritised for their relevance to prematurity: fetal anomalies, intrauterine growth restriction (IUGR), prematurity and stillbirth gene sets. We observed significant differences in the proportion of term, moderately preterm, and very or extremely preterm probands with diagnostic mutations in the fetal anomalies gene set (p=3.33 x 10^-5^) (**Supplementary Table 8**). Post-hoc analysis indicated that moderately preterm probands had a significantly higher enrichment of mutations in the fetal anomalies gene set compared to term probands (OR=1.36, p=7.83×10^-5^), while very or extremely preterm probands exhibited a nominal enrichment of these mutations compared to term probands (OR=1.54, p=1.92×10^-2^) (**Figure 3**). We saw a nominal enrichment of diagnostic mutations in premature probands in the gene sets associated with IUGR and prematurity (2.11×10^-2^ < p < 0.05)(**Figure 3**). Post-hoc comparisons demonstrated a nominal difference in the proportion of moderately preterm probands with mutations in these gene sets when compared to term probands (8.53×10^-3^ < p < 0.05)(**Figure 3**).

**Figure 3.**
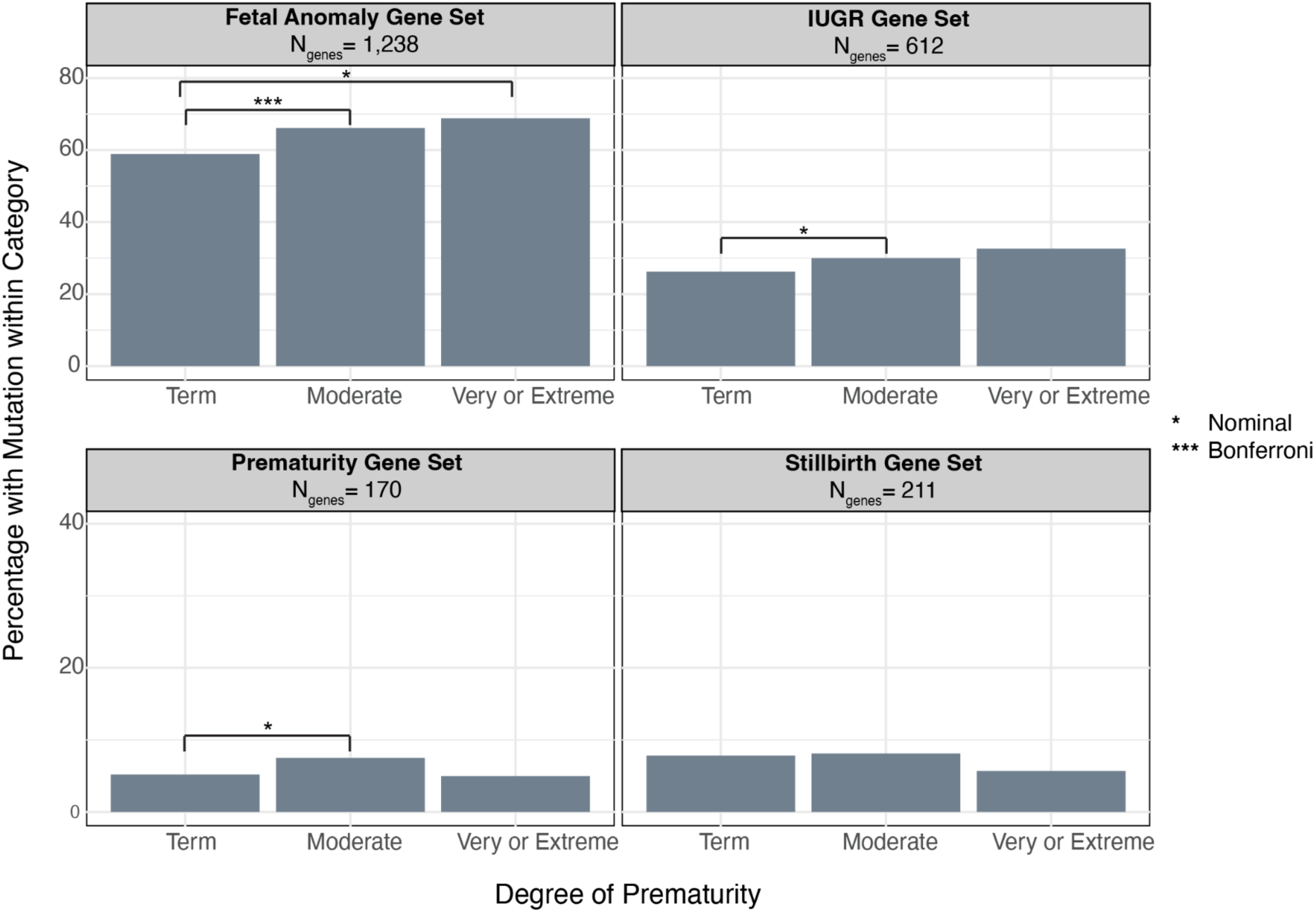
Gene set enrichment analysis. Bar chart showing the proportion of diagnosed probands within each gestational age group with a diagnostic mutation in gene from a given gene set. The fetal anomaly^28^ and stillbirth^29^ gene sets are curated by experts. Gene sets associated with prematurity and IUGR were derived by the HPO team from a combination of OMIM data mining and manual curation.^30^

In summary, we find some evidence that monogenic diagnoses in certain genes and gene sets are differentially prevalent across term and preterm probands in this sample. However, these enrichments could have several possible interpretations, as discussed below.

### The relationship between *de novo* variation and gestational age in developmental disorders

As *de novo* mutations (DNMs) account for the majority of genetic diagnoses among DDs,^5,31^ we sought to compare the fraction of diagnoses attributable to DNMs and the genes enriched for DNMs between preterm (N=2,169) and term (N=11,857). Overall, both preterm and term groups carried significantly more nonsynonymous DNMs than expected under a null mutational model (p<4.38×10^-63^)(**Figure 4**; **Supplementary Table 9**). We previously observed that term probands were more likely to have a genetic diagnosis than preterm probands in DDD^5^, reflecting a liability threshold model; consistent with this, the attributable fraction across all genes was nominally significantly higher for term probands than preterm probands (39% [95% CI: 37-41%] versus 33% [29-37%]; Z-test p =0.01)(**Figure 4**; **Supplementary Table 10**).

**Figure 4.**
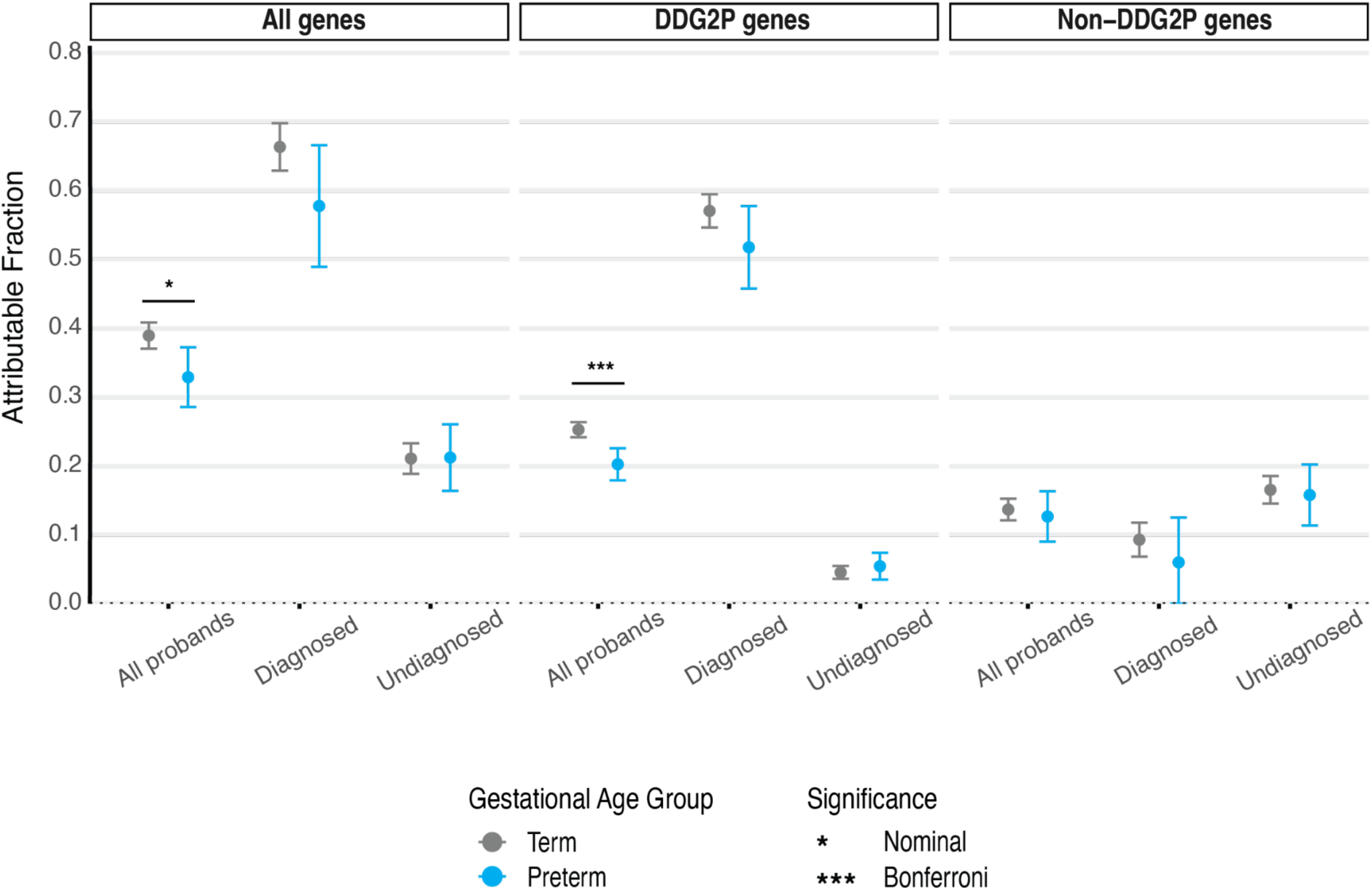
Attributable fraction of preterm and term developmental disorder cases due to *de novo* variation. The y-axis depicts the attributable fraction of cases accounted for by nonsynonymous *de novo* mutations, shown separately for term and preterm probands, both overall and stratified by diagnostic status. The figure presents the attributable fraction across all genes as well as for genes with and without known DD associations (DDG2P and non-DDG2P, respectively). Asterisks indicate the significance of Z-tests comparing the fraction between each group of term and preterm probands. Error bars represent 95% confidence intervals.

We then further compared attributable fractions between DD-associated genes and all other genes. Term infants had a higher attributable fraction than preterm among known DD-associated genes (25% [95% CI:24-26%] versus 20% [95% CI:18-23%]; Z-test p =1.28×10^-4^) but there was no difference between these groups in genes with no known DD-associations (14% [95% CI:12-15%] versus preterm 13% [95% CI:9-16%]; Z-test p =0.62) (**Figure 4**). Among undiagnosed probands across all genes, DDG2P genes and non-DDG2P genes, there was no difference in the attributable fraction between the term and preterm probands, but both groups were enriched for non-synonymous mutations (p<1.68×10^-10^) (**Supplementary Table 10**). Overall, these findings suggest that while premature probands are less likely to have genetic diagnoses than term probands in these cohorts, there are still diagnoses to be made in both known and novel DD-associated genes among both preterm and term undiagnosed probands.

### Characterising the association between polygenic scores for educational attainment and gestational duration

Since genetic variation may confound associations between epidemiological factors such as prematurity and clinical outcomes in DDs, in this work, we aimed to investigate whether genetic background influences the relationship between gestational age at birth and phenotypic outcomes in DDs. Prior to testing this hypothesis, we first sought to replicate and further explore associations between common variant predisposition to education-related traits and prematurity that were observed in probands with neurodevelopmental conditions in DDD^3^, across DDD probands (N=7,032) and DDD-like probands from 100kGP (N=4,781). We considered polygenic scores (PGSs) for three education-related traits: educational attainment (PGS_EA_)^32^, and its cognitive (PGS_CogEA_) and non-cognitive components (PGS_NonCogEA_)^33^. The non-cognitive component captures factors such as personality traits and socioeconomic status that may influence educational attainment independently of cognitive ability,^33^ and is relevant to this study as some of these factors are correlated with gestational duration.^34–38^

Associations between the child’s gestational age at birth and their PGS varied between the cohorts; in DDD, they were consistent with observed genetic correlations^3^ and were generally stronger than in 100kGP, possibly due to the larger sample size (**Supplementary Figure 7**). In the cross-cohort meta-analysis of all probands, gestational duration was nominally significantly associated with the PGS_EA_ (*β*=0.02, p=0.03) and significantly associated with PGS_NonCogEA_ (*β*=0.03, p=1.83х10^-3^). This suggests that probands with a higher common variant predisposition to the non-cognitive components of EA were more likely to be born at later gestational ages (**Figure 5**). When stratifying the probands by diagnostic status, we only observed an association between PGS_NonCogEA_ and gestational duration in undiagnosed probands (*β*=0.05, p=6.82х10^-5^; also nominally significant in both cohorts), implying that the presence of a diagnostic variant attenuates the effect of PGS_NonCogEA_ on gestational duration (**Figure 5**). No association was observed between the number of inherited damaging coding variants in loss-of-function-intolerant genes, captured as a rare variant burden score (RVBS), and gestational duration, although these rare variants are also known to be associated with lower EA^39–42^. As a sanity check, we tested whether PGS for gestational duration^14^ was associated with this phenotype, and found that it was in the meta-analysis of all probands (*β*=0.02, p=1.62х10^-2^) (**Figure 5**).

**Figure 5.**
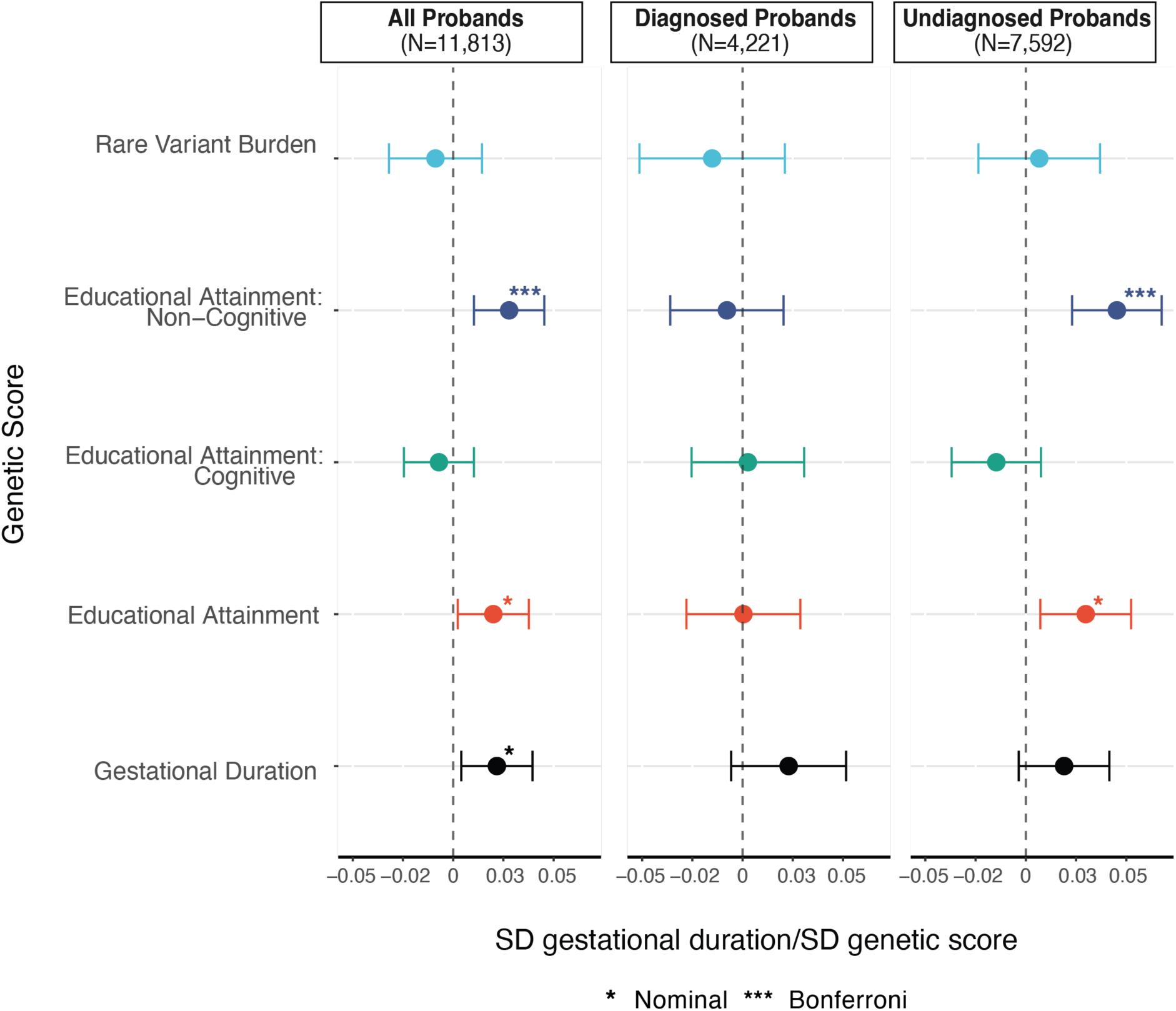
Associations between genetic measures and gestational duration. Standardised effect of proband’s genetic measure on gestational duration. Gestational duration was rank-based inverse normal transformed (RINT). The effect size should be interpreted as change in standard deviation of RINT gestational duration per standard deviation change in genetic score. Panels show effect size estimates from a meta-analysis of regression results from DDD and 100kGP. Error bars represent 95% confidence intervals.

We next tested whether associations between a proband’s PGS for education-related traits and gestational duration are driven by direct genetic effects and whether there was any evidence for these associations being driven by parental alleles which presumably act by affecting the prenatal environment (ie. indirect genetic effects). To test this, we fitted a trio model^43^ in which we regressed the child’s gestational age at birth on their own PGS plus the PGSs of their parents. In this model, the coefficient on the child’s PGS represents the direct genetic effect, whereas the coefficient on the parents’ PGSs represents the association between non-transmitted parental alleles and the child’s phenotype, which could reflect either indirect genetic effects or confounders.^43–45^ We found no evidence of a direct genetic effect of probands’ PGS_NonCogEA_ on gestational duration (**Supplementary Figure 8**). In contrast, we detected a nominal significant positive association between maternal and paternal non-transmitted alleles for PGS_NonCogEA_ and PGS_EA_ and gestational duration. These findings suggest that the observed association between the proband’s PGS for EA and its non-cognitive component may reflect the influence of parental polygenic predisposition to these traits on gestational duration, rather than a direct effect of the proband’s genotype. However, we cannot rule out that this is driven by confounders such as parental assortment or uncontrolled population stratification.^3,43,45,46^ (See <u>Supplementary Note 3</u> for further discussion of the trio analysis.)

Finally, we assessed whether the observed association between PGS_NonCogEA_ and prematurity might confound the relationship between prematurity and clinical outcomes in DDs. We repeated the phenotype association analysis while controlling for PGS_NonCogEA_ and observed no significant change in the associations between degree of prematurity and any antenatal factors or tested phenotypic outcomes in either cohort (**Supplementary Table 11**). Given that PGS_NonCogEA_ may capture aspects of socioeconomic status, we conducted a similar analysis adjusting for the Index of Multiple Deprivation^47^, a measure of socioeconomic status available for 100kGP probands. Again, we observed no significant change in associations (**Supplementary Table 12**). Therefore, the association between prematurity and these phenotypes is unlikely to be confounded by socioeconomic status or common variants associated with the non-cognitive component of EA.

## Discussion

Using data from two large cohorts of individuals with DDs recruited for clinical sequencing, we investigated genetic influences on premature birth, and associations between prematurity and phenotypic outcomes. We demonstrated that prematurity, in addition to the presence of a monogenic diagnosis, impacts the clinical features and developmental trajectories of these individuals. We identified developmental disorder genes and relevant gene sets associated with an elevated risk of preterm delivery among individuals with DDs, motivating future research aimed at understanding the relationship between prematurity and genetic variation in DDs. Our analysis of *de novo* variation demonstrated that a higher proportion of probands are attributable to DNMs in known disease-genes than in preterm probands, but for both groups there is a similar proportion of probands accounted for by DNMs in as-yet-undiscovered developmental disorder genes. Finally, we show that individuals with higher common variant predisposition to the ‘non-cognitive’ component of EA are, on average, less likely to be born premature, but that this association is not driven by direct genetic effects on the child.

Our work shows that prematurity expands the spectrum of clinical features experienced by individuals with DDs ascertained for clinical sequencing. Specifically, prematurity was associated with an increased number of affected organ systems. We identified organ systems related to common complications of prematurity — in particular respiratory,^48^ cardiovascular^49^ and digestive systems^48^ — to be more frequently affected among premature probands; this held even after accounting for the presence of a monogenic diagnosis (**Figure 1**). These findings align with recent work in autism^13^ which showed that individuals born prematurely had more comorbidities than those born at term. Although both prematurity and the presence of a monogenic diagnosis^5^ are associated with neurodevelopmental impairment,^50–58^ we did not find an increased rate of neurodevelopmental abnormalities among premature probands in our analysis of HPO terms.^9,59^ We suspect this discrepancy may reflect cohort ascertainment: nearly all probands have an HPO term related to abnormal neurodevelopment, reducing our power to see any effect of prematurity on this. However, our milestone analysis does provide support for an additive impact of prematurity and monogenic diagnoses on neurodevelopment. Premature probands with a monogenic diagnosis experience greater delays than those without a monogenic diagnosis in attaining certain developmental milestones, including sitting and walking independently and first words (**Figure 2**). Deeper and longitudinal phenotyping, such as through integration of electronic data on educational records,^60,61^ will be required to partition the relative contributions of environment and genetics on longer-term neurodevelopmental outcomes.

For our analysis of the role of rare genetic variation, we hypothesised that premature individuals with DDs may have proportionally more diagnostic variants in genes with links to fetal anomalies, IUGR, prematurity and stillbirth than term individuals with DDs. We found an enrichment of diagnostic variants in genes from the fetal anomalies gene panel in moderate and very/extremely preterm individuals compared to term individuals (**Figure 3**). This is unsurprising as congenital anomalies are found at an increased rate among individuals born premature.^62^ However, there was no significant enrichment of diagnostic variants in genes linked to IUGR, prematurity and stillbirth among preterm individuals. This may reflect the lack of a robust knowledge base of the rare genetic causes of these traits.^15,29^ These latter three lists are also smaller, reducing our power.

We then performed an analysis of odds of prematurity across individual disease genes in which 30 or more diagnostic variants were observed in our cohorts. Although we had adequate power to detect genes with an odds ratio of six or higher, no genes remained significant after correction for multiple testing, suggesting that the most prevalent monogenic causes of DDs are unlikely to have a very high impact on the risk of premature birth among individuals already ascertained for DDs. At a nominal level of significance, we identified *EP300*, *KMT2A*, *NSD1,* and *NF1* as increasing the likelihood of premature birth and *MECP2* as decreasing it. The association with EP300 reached statistical significance in a sensitivity analysis restricted to individuals with a clinically confirmed pathogenic variant. Fetuses with mutations in *EP300* are predisposed to IUGR, and their mothers are at increased risk of preeclampsia,^63^ both of which are indications for medical induction of preterm labour. Given this, there is a case for reconsidering the comment in the recent consensus guideline for Rubinstein-Taybi syndrome stating there is no increased risk of premature birth for individuals with variants in *EP300*.^64^ In future, large datasets of preterm children, ideally ascertained for being born preterm rather than for having a DD, will be required to clarify the association between the genes identified in our study and prematurity risk.

Our analysis of the burden of *de novo* variation lends further support to a liability threshold model of DD aetiology and has potential clinical implications. We found that the attributable fraction of cases to nonsynonymous DNMs among all genes was nominally higher for term than preterm probands, and that among DDG2P genes alone this difference was significant after correcting for multiple testing (**Figure 4**). This is consistent with our previous findings which considered the rate of genetic diagnoses, and with a model in which an individual’s total liability must exceed a certain threshold to manifest clinically recognisable DDs: those with a substantial environmental contribution require less or no genetic contribution.^5^ This finding suggests that initial clinical genome sequencing analysing known disease genes will generally yield more *de novo* diagnoses if focused on term probands, whilst novel gene discovery efforts are likely to be equally helpful for diagnosing preterm and term probands. Interestingly, no difference in the rate of DNMs in neurodevelopmental disorder genes was observed between term and preterm probands in autism^13^, perhaps due to its different genetic architecture and overall lower contribution from DNMs compared to the DDs studied here.

This work advances current understanding of the contribution of common genetic variation to preterm delivery in DDs two key ways. First, we demonstrate that while common variants associated with the non-cognitive component of EA are positively associated with gestational age at birth across all DD probands, this association becomes null when the analysis is restricted to probands with a monogenic diagnosis (**Figure 5**). This finding suggests that among diagnosed probands, it is the nature of the genetic diagnosis that has the largest effect on gestational timing, and this partly overrides the effect of any polygenic factors that might otherwise influence the likelihood of premature birth. Second, we show that the association between PGS_NonCogEA_ and prematurity in probands is not explained by direct genetic effects **Supplementary Figure 8**). Instead, parental non-transmitted alleles associated with the non-cognitive component of educational attainment are associated with the risk of preterm birth, likely due to effects on the prenatal environment or confounding factors like population stratification or assortative mating. Non-cognitive skills associated with higher EA, such as self-regulation and motivation, are negatively genetically correlated with substance use, risk-taking behaviours, and socioeconomic deprivation.^33^ Maternal smoking and drug use have been identified as potential mediators of the established epidemiological association between maternal educational attainment and preterm delivery.^35^ Given this context, our trio analysis supports the idea that parental behaviours associated with the non-cognitive component of educational attainment may contribute to the risk of preterm birth in offspring.

Our analysis has several limitations. First, we use phenotypic terms recorded on assessment for analyses; the selection of terms is dependent upon the clinician and phenotyping protocols. Indeed, the number of individuals with terms related to several prematurity phenotypes is below what would be expected, and this limits our ability to detect associations. For example, the prevalence of retinopathy of prematurity among babies with a birthweight of less than 1500g is 12.5% in the UK^65^ and yet, with 790 such individuals across the two studies, no individuals have a recorded HPO term related to this condition. This may be because clinicians focused on recording terms they thought likely to be associated with monogenic diseases. Relatedly, when we attempted to correct for gestational duration in the milestone analysis, this highlighted inconsistencies that suggested that the milestones for some but not all probands had been corrected for gestational age before data entry (see Supplementary Note 1), which likely added noise in our analyses. Another limitation is that the rare variant analysis is performed in individuals with DDs; thus, it is not informative about the effect of rare variants in a given gene on the unconditional chance of premature birth within the general population. Instead, it simply indicates whether variants in specific DD-associated genes are more likely to be seen in premature probands amongst this sample of DD patients recruited for clinical sequencing. Indeed, premature probands are only likely to have been recruited if their clinical phenotype was disproportionate to the expected effects of their prematurity. This clinical threshold for inclusion could introduce correlations between variables (e.g. a specific monogenic syndrome and prematurity) that would not exist in a random, unascertained sample (i.e. collider bias).

In summary, we begin to tease apart the relative contributions and interplay of common and rare genetic variation and prematurity in developmental disorders. In future, the genetic and phenotypic characterisation of research cohorts ascertained for being born prematurely would provide more generalisable findings. Such cohorts would enable us to establish the fraction of premature babies likely to have a monogenic diagnosis, identify rare variants which increase the risk of being born prematurely, and investigate which clinical features indicate that a premature baby is likely to benefit from genetic investigations for a monogenic cause. Emphasis should be placed on sampling those at the lower extremes of gestational duration, who are most likely to experience health sequelae but are uncommon in the general population. This would allow us to understand the complex relationships between gestational duration, genetic variation, and health and development.

## Methods

### Sample Overview

#### Deciphering Developmental Disorders (DDD)

This study includes 13,401 probands from the DDD study, which aimed to identify molecular diagnoses for families and patients with previously genetically undiagnosed DDs that were thought likely to be of genetic origin.^1^ Enrolment occurred between 2011 and 2015 from 24 clinical genetics units in the United Kingdom and Ireland. The DDD cohort is described in detail elsewhere.^1,5,66,67^

#### Genomics England 100,000 Genomes Project (100kGP)

This study uses data from the rare disease programme of the 100,000 Genomes project, an initiative by the UK Department of Health and Social Care to whole-genome sequence individuals from the National Health Service with rare conditions and cancer.^68,69^ We restricted our analysis to probands with phenotypes similar to those recruited into DDD with information on gestational duration (DDD-like, n=9,310). DDD-like probands from the 100,000 Genomes projects are defined as individuals who:

1. were recruited into the same disease model as GEL probands who had previously been enrolled in DDD, or
2. had one of the top five HPO terms used in DDD and their descendants, including HP:0000729 (autistic behaviour), HP:0001250 (seizure), HP:0000252 (microcephaly), HP:0000750 (delayed speech and language development) and HP:0001263 (global developmental delay).

Probands with an age of onset >16 years were excluded, as well as probands recruited into the neurodegenerative disorders subcategory or recruited into a disease subcategory for which the mean age of probands was >16 years. Lastly, we excluded any participants who were also recruited into DDD and/or who were related to DDD participants, leaving 8,311 probands for analysis.

#### Description of Phenotypes

For DDD probands, demographic, gestational duration, birthweight and HPO^30^ terms were recorded by the recruiting clinical geneticists in DECIPHER^70^ and additional phenotypic information including on developmental milestones was collected using a bespoke online questionnaire collected via DECIPHER (**Supplementary Table 1**). See <u>Supplementary Note</u> <u>1</u> for a description of developmental milestone data and quality control procedures.

For 100kGP probands, HPO terms were recorded by the recruiting clinicians. Gestational duration and birthweight data in 100kGP was gathered for all rare disease probands using three data sources (**Supplementary Figure 9**). First, about 1000 probands had this data clinically ascertained during recruitment. Second, the 100kGP has linked electronic healthcare data including admitted patient care episodes. Gestation and birthweight are recorded at birth in this dataset in the ‘gestat’ and ‘birtweit’ columns in episodes relating to both the child and mother, providing two potential data sources.^71^ The z-scores for birthweight were then calculated using “British 1990 reference data, reanalysed 2009”.^72^ Individuals with a birthweight more than 5 standard deviations from the mean for their gestational duration and sex were removed (**Supplementary** Figure 10-11). The data was then limited to DDD-like probands (N=8,311). This data is available to share within the Genomics England Research Environment upon request.

Preterm birth was defined as delivery before 37 completed weeks of gestation and was further categorised according to the World Health Organisation definitions.^8,21^ We selected antenatal factors available in both cohorts (including abnormalities on antenatal ultrasound scan, multiple pregnancy, maternal diabetes during pregnancy, and a history of previous pregnancy loss), birthweight and HPO terms for analysis. We also considered a list of medically-relevant HPO terms curated for their relevance to DDs^2^, including abnormal speech or language, any craniofacial cleft, atypical behaviour, hearing or visual impairment, hypotonia, abnormal head circumference, seizure and short stature. We counted the number of probands with an abnormality of an organ system or organ and with medically-relevant HPO terms by conducting a search of a particular term and its daughter nodes.^73^ We determined the number of distinct affected organ systems in each proband, following the methodology outlined in Niemi *et al*.^2^, which accounts for overlapping terms to avoid redundancy across multiple organ systems. **Supplementary Table 1** provides an overview of the phenotypic measures from DDD and 100kGP included in this study.

### Genetic Data Preparation

#### DDD

DDD probands were genotyped using the Illumina HumanCoreExome chip (CoreExome), the Illumina OmniChipExpress (OmniChip), and the Illumina Infinium Global Screening Array (GSA). A subset of probands were genotyped using more than one array. For this study, we used CoreExome and OmniChip data for analysis of the probands and the GSA and OmniChip data for analyses involving trios. Quality control procedures for the genotype data are described in detail elsewhere.^2,3^ WES data was used for the analysis of rare variants. Quality control and processing of the exome sequencing data are described in Huang *et al*.^3^

#### 100kGP

Whole genome sequencing of 100kGP participants was performed with 150bp paired-end reads using Illumina HiSeqX. This study used the 78,195 germline genomes from the Aggregated Variant Calls released by the 100kGP team. Quality control and processing of the whole-genome sequencing data are described in Huang *et al*.^3^

#### Genetic ancestry inference

We restricted our analysis to individuals of genetically-inferred British ancestry (GBR-ancestry) defined by genetic similarity to British individuals from the 1000 Genomes Project.^74^ The process of identifying GBR-ancestry samples in DDD and GEL was described previously ^2,3^. Briefly, a set of LD-pruned overlapping SNPs from DDD and 100kGP with MAF >5% from a subset of unrelated individuals were projected onto 1,000 Genomes phase 3 individuals^74^. Another principal component analysis was performed with the European ancestry subset and a homogeneous subgroup with GBR-ancestry was identified for each cohort.

#### Identification of related participants

We used KING^75^ to identify up to third-degree relatives (kinship coefficient > 0.0442) in each cohort and used a subset of unrelated individuals (i.e. those more distantly related than third-degree) for genetic analyses (N_DDD_=7,052; N_100kGP_=4,781). For analyses of trios, we further restricted our analysis to probands whose parents were unrelated to other parents in the cohort (N_DDD trios_=3,101; N_100kGP trios_=2,989).

#### Genotype imputation

For DDD probands, imputation was performed separately for samples from each genotype array using the maximum number of available variants after QC and removal of palindromic SNPs. The HRC r1.1 reference panel^76^ was used for imputation of samples genotyped on the CoreExome array^2^ and the TOPMed r2 reference panel^77^ was used for imputation of GSA and Omnichip samples^3^. Well imputed SNPs (Minimac4 R^2^ >0.8) with MAF >1% present in the GEL 100kGP data were retained for further analysis (N=5,699,435).

#### De novo variant calling

DNM calling and quality control procedures for 9,858 trios from the DDD study are described in detail elsewhere.^78^ For this analysis, DNM calling was carried out for 13,381 trios from 100kGP following the same quality control procedures, 5,991 trios met the inclusion criteria for this study (Supplementary Methods).

### Defining a monogenic diagnosis

#### DDD

Potentially clinically relevant rare variants were identified from WES data and chromosome microarray data using a variant analysis pipeline described in Wright et al.^5^ Briefly, the study team identified rare damaging variants that fit an appropriate inheritance pattern from a set of genes known to be causal for DDs (DDG2P, https://www.deciphergenomics.org/ddd/ddgenes). Candidate diagnostic variants were uploaded to DECIPHER^70^ and the pathogenicity was annotated by the patients’ referring clinician. “Diagnosed” probands were defined as those with at least one variant with a clinical annotation of pathogenic/likely pathogenic in DECIPHER, or with a predicted classification of pathogenic/likely pathogenic using autocoded ACMG diagnoses.^5^ All remaining probands were classified as “undiagnosed”.

#### 100kGP

For the 100kGP genomes, diagnostic rare variant and copy number tiering was performed as previously described.^79,80^ We categorised the diagnostic status of all probands included in the Genomic Medicine Service exit questionnaire. This questionnaire required the referring clinician to classify the pathogenicity of candidate variants identified by 100kGP’s variant analysis pipeline and an overall diagnostic classification for the family. We defined “diagnosed” probands as those with a variant that is annotated as pathogenic or likely pathogenic and the family classified as being ‘solved’ or ‘partially solved’ by the referring clinician. We also classified probands who have a variant prioritised by the 100kGP Clinical Research Team, and awaiting clinical review, in the ‘Diagnostic Discovery data’ as “diagnosed” due to the high reported probability of these variants being later classified as disease-causing.^81^ All other probands were classified as “undiagnosed”.

### Calculation of Polygenic Scores

Polygenic scores (PGS) were calculated using summary statistics from the latest genome-wide association studies (GWAS) for gestational duration^14^, EA^32^ and the cognitive and non-cognitive components of EA^33^. PGS were estimated using LDpred2-auto^82^ for a set of of 1,444,196 HapMap3+ variants^83^ that pass quality control in 100kGP aggV2 samples, have a minor allele frequency (MAF) > 1%, and are well-imputed in all genotyping arrays used in DDD (Minimac4 R^2^ > 0.8). The recommended LD reference panel generated using HapMap3+^83^ variants was used for PGS calculations. PGS for each individual were estimated using PLINK v1.9 ^84^, which uses the weights generated by LDpred2 to calculate the weighted sum of genotypes across a set of SNPs for each individual.

### Calculation of Rare Variant Burden Scores

Sequence data from parents and probands from DDD and 100kGP was annotated with the Ensembl Variant Effect Predictor (VEP)^85^ and variants with the “worst consequence” annotation across transcripts were retained. Autosomal heterozygous protein-truncating variants (PTVs) defined as high confidence by LOFTEE^86^ and “missense” variants (defined as missense, stop lost, start lost, inframe insertion, inframe deletion, and loss-of-function variants annotated as low-confidence by LOFTEE^86^) were extracted. Rare variants with a MAF < 1 x 10^-5^ in each gnomAD super-population and MAF < 1 x 10^-4^ in the respective cohorts were retained for analysis. A deleterious rare variant burden score (RVBS) was calculated by summing the number of high confidence PTVs and missense variants (MPC ≥ 2^87^) in loss-of-function-intolerant genes (pLI > 0.9), restricting to inherited variants in children.

### Data Analysis

All data analysis was performed using R v4.3.1^88^ unless otherwise specified. The analysis plan was pre-registered^89^ and deviations from the analysis plan are described in the <u>Supplementary</u> <u>Methods</u>.

#### Associations between prematurity and phenotypic outcomes

We used multivariable logistic regression models to assess the association between degree of prematurity and binary phenotypic measures listed in **Supplementary Table 1**, and multivariable linear regression models for continuous phenotypic measures, excluding developmental milestone attainment (see <u>time-to-event analysis</u>). Each degree of prematurity, “moderate”, “very” or “extreme”, was compared to being born at term. The baseline models were estimated in the entire sample and regressed the phenotypic measure on degree of prematurity while controlling for relevant covariates.

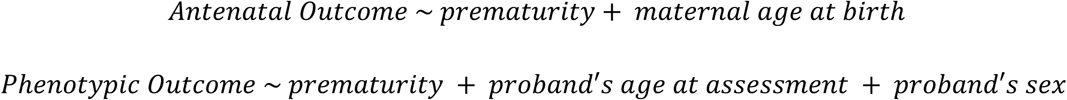

where [*phenotypic outcome*] refers to all other phenotypic measures besides the antenatal outcomes listed in **Supplementary Table 1.**

To test if the presence of a diagnostic variant modified the association between prematurity and neurodevelopmental outcomes, an additional set of models was fitted to adjust for genetic diagnosis and an interaction between prematurity and a genetic diagnosis.

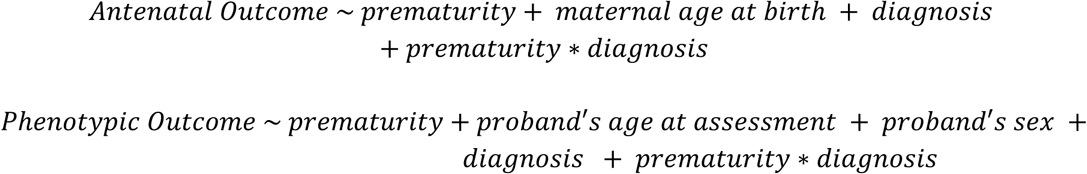

Finally, to assess the influence of other factors on the relationship between prematurity and phenotypic outcomes, we re-estimated all models, each time including only one additional factor. Specifically, we adjusted for either a deprivation index (available in 100kGP only) or genetic scores that showed a significant association with gestational duration.

We applied the Bonferroni correction for 522 tests (29 phenotypes x 3 levels of prematurity x 6 models per phenotype) and considered p<9.58×10^-5^ significant. Regression models were estimated in DDD and 100kGP separately; and effect size estimates were meta-analysed using an inverse-variance weighted approach for phenotypes assessed in both cohorts.

#### Time-to-event analysis

Kaplan-Meier survival analysis with log-rank tests was performed to test for differences in the time taken to achieve a developmental milestone (social smile, sitting independently, walking independently, and first words) across gestational age groups. Analysis was performed for a subsample of participants from the DDD study with data on developmental milestone attainment (*n*=5,692-11,726). We also estimated Cox proportional hazards for developmental milestone attainment for each category of prematurity, relative to being born at term. The Cox models were adjusted for proband’s age, sex, genetic diagnostic status, and an interaction between genetic diagnosis and degree of prematurity. We applied the Bonferroni correction for multiple testing, p<0.05/4 (1 log-rank test for 4 milestones) and p<0.05/24 ([3 degrees of prematurity + 3 interaction terms] for 4 milestones) were considered significant for Kaplan Meier analysis and Cox models respectively.

#### Testing for enrichment of monogenic diagnoses in specific genes and gene sets

Logistic regression analyses were used to test for an association between prematurity, encoded as a binary variable, and the presence of monogenic diagnoses in selected genes in diagnosed probands from DDD and 100kGP. Based on power calculations, we conducted association testing for 44 genes with at least 30 observed diagnostic variants in our dataset (<u>Supplementary Methods</u>) and applied Bonferroni correction to account for the 44 tests.

Fisher’s exact tests with post-hoc pairwise comparisons were conducted to test for an enrichment of monogenic diagnoses in four gene sets associated with adverse pregnancy outcomes (fetal anomalies, prematurity, stillbirth, and intrauterine growth restriction) (<u>Supplementary Methods</u>) across the gestational age groups in diagnosed probands from DDD and 100kGP. Tests that pass the Bonferroni correction (α = 0.05/(4 gene sets tested × 4 tests per gene set); p < 3.13 × 10^-3^) were considered significant.

#### Estimating excess DNMs and the attributable fraction

We compared the observed number of DNMs to the expected count predicted by a null mutational model^90^ for each consequence class, across the exome as well as in genes with and without a known association with DDs according to DDG2P^91^. We conducted Poisson tests to assess differences in the observed and expected nonsynonymous DNM counts and applied a Bonferroni multiple testing correction to account for 18 tests (2 gestational age groups x 3 proband groups x 3 gene sets).

We also estimated the attributable fraction, the proportion of DD cases attributable to nonsynonynous DNMs using the following formula:

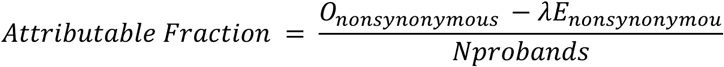

Where *O_DMN_* is the observed number of nonsynonynous DNMs, *E_DMN_* is the expected number calculated using the model from ^90^, and λ is a correction factor calculated as 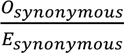 among all probands.

The attributable fraction was estimated for probands in each gestational age group, both overall and stratified by diagnostic status, across the exome and for genes with and without known DD associations^91^. We compared the attributable fraction between gestational age groups for each gene set using Z-tests and applied a Bonferroni correction to account for nine tests (3 proband groups x 3 gene sets).

#### Associations between genetic measures and gestational duration

We tested for an association between gestational duration and four PGS (gestational duration and three PGS for educational attainment) as well as RVBS by fitting linear regression models in unrelated probands of genetically inferred GBR ancestry. Since gestational duration was non-normally distributed, we applied a rank-based inverse normal transformation. Proband’s sex and the first 20 genetic principal components (PCs) were included as covariates in the models. We ran the model using data from all probands, as well as separately for diagnosed and undiagnosed probands.

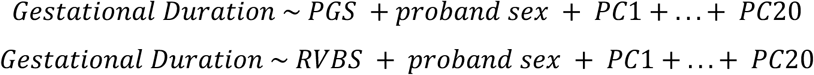

We applied a Bonferroni correction for fifteen tests (five genetic measures and three proband groups - all, diagnosed, and undiagnosed probands). All models were estimated in DDD and 100kGP100k separately. The effect size estimates from the two cohorts were meta-analysed using an inverse-variance weighted approach.

Lastly, we extended the baseline model in fully genotyped trios to control for parental PGS to test if maternal and paternal non-transmitted common alleles are associated with offspring gestational duration (Supplementary Methods).

## Data Availability

The raw and post-QC genotype array data and exome sequence data from DDD are available through European Genome-phenome Archive, under <u>EGAS00001000775</u>. Whole-genome sequence data and phenotypic data from the 100,000 Genomes project can be accessed by application to Genomics England (https://www.genomicsengland.co.uk/research/academic/join-gecip). Birthweight and gestational age data, and relevant code, is available upon request in the 100kGP for registered users.

## Acknowledgements

We are grateful to the families whose participation in the DDD study and the 100,000 Genomes Project made this research possible. We also extend our gratitude to our colleagues who assisted in the generation and processing of data, including teams at the DDD project and Genomics England. We also thank Daniel Malawsky and Helen Firth for helpful discussions.

DDD: The DDD study presents independent research commissioned by the Health Innovation Challenge Fund (grant no. HICF-1009-003). See www.ddduk.org/access.html for full acknowledgements. This study makes use of DECIPHER, which is funded by the Wellcome Trust.

10,000 Genomes Project: This research was made possible through access to data in the National Genomic Research Library, which is managed by Genomics England Limited (a wholly owned company of the Department of Health and Social Care). The National Genomic Research Library holds data provided by patients and collected by the NHS as part of their care and data collected as part of their participation in research. The National Genomic Research Library is funded by the National Institute for Health Research and NHS England. The Wellcome Trust, Cancer Research UK and the Medical Research Council have also funded research infrastructure.

## Funding Statements

The DDD study presents independent research commissioned by the Health Innovation Challenge Fund (grant number HICF-1009-003). This study makes use of DECIPHER, which is funded by the Wellcome Trust. The full acknowledgements can be found at https://www.ddduk.org/accessing-ddd-data/. This research was funded in whole or in part by the Wellcome Trust Grant 220540/Z/20/A, ‘Wellcome Sanger Institute Quinquennial Review 2021–2026’, Wellcome Trust Grant 226083/Z/22/Z. For the purpose of open access, the authors have applied a CC-BY public copyright license to any author accepted manuscript version arising from this submission.

PC was funded by King’s College London EPSRC Centre for Doctoral Training in DRIVE-Health, Grant Ref: EP/Y035216/1.

## Author Contributions

OW and PC conducted most of the analyses, with the remainder being conducted by SJL and ED. QH, PC, OW, SA, and SJL carried out data preparation and quality control with supervision by HCM. EJR and HCM supervised the analyses and directed the study with key intellectual input from SR and MAS. OW, PC, EJR and HCM wrote the first draft of the manuscript, with input from SR. All authors read and commented on the final manuscript.

## Ethics Declaration

All data were collected with informed consent and in accordance with relevant guidelines and regulations. The DDD study is approved by the UK Research Ethics Committee (10/H0305/83, granted by the Cambridge South Research Ethics Committee and GEN/284/12, granted by the Republic of Ireland Research Ethics Committee). The 100,000 Genomes project was approved by the East of England-Cambridge Central Research Ethics Committee (REF 20/EE/0035).

## Competing Interests

MEH is a co-founder of, consultant to and holds shares in Congenica, a genetics diagnostic company, and is also a consultant to AstraZeneca Centre for Genomics Research. All other authors declare no competing interests.

