## Supplementary Information for "Investigating the Interplay Between Prematurity and Genetic Variation in the Context of Rare Developmental Disorders"

#### Supplementary Methods

##### *De novo* mutation filtering in 100,000 Genomes Project

We analysed *de novo* mutations (DNMs) in 5,991 trios from the 100,000 genomes project (100kGP). DNMs were called using the Platypus variant caller[1] by the Genomics England Bioinformatics team. The following filters were applied to both cohorts (100kGP and DDD):

- Removal of DNMs in non-coding regions
- Removal of DNMs within known segmental duplication regions defined by UCSC (http://humanparalogy.gs.washington.edu/build37/data/GRCh37GenomicSuperDup.tab)
- Removal of DNMs in highly repetitive regions (http://humanparalogy.gs.washington.edu/build37/data/GRCh37simpleRepeat.txt)
- Retain only the most severe DNM per gene for each individual based on consequence severity as determined by VEP[2]
- Removal of indels > 100bp
- Removal of variants with allele frequency in gnomAD > 0.01
- Removal of samples with > 10 DNMs
- Randomly retain one DNM per sibling pair if DNM is present in both siblings

The filters applied specifically to 100kGP were as follows:

- Genotype is heterozygous in child (1/0) and homozygous in both parents (0/0)
- Read depth > 20
- Child read depth < 98[3]
- VAF > 0.3
- Removed DNMs present in unaffected probands

##### Power to detect enrichment of monogenic diagnoses in specific genes

To estimate the power to detect enrichment of monogenic diagnoses in specific genes, we simulated datasets consisting of 984 preterm probands and 6,462 term probands (i.e. the number of diagnosed probands with clinically confirmed diagnoses in DDD and GEL) with the frequency of observed mutations in a gene ranging from 10-55 and the odds ratio for a mutation being associated with prematurity varying from 2-10. We simulated 10,000 datasets for each combination of odds ratio and mutation rate and fit a logistic regression model to test for an association between prematurity and having a diagnosis for each simulated dataset. We calculated the power to detect an association between a diagnostic mutation in a specific gene and prematurity as the proportion of significant p-values out of 10,000 null datasets, adjusted for the number of genes tested, for each combination of odds ratio and mutation rate.

We estimate that for mutations with an odds ratio of ≥ 6, we have 80% power to detect an enrichment of monogenic diagnoses in premature probands in genes for which at least 30 mutations are observed in our dataset.

#### Definition of gene sets

We compared the proportion of probands in each gestational duration group (term, moderately preterm, and very or extremely preterm) with genetic diagnoses in the following gene sets:

1) A set of 1,239 genes with a disease confidence rating of "confirmed" from The NHS Genomics Medicine Fetal Anomalies panel curated by the Prenatal Assessment of Genomes and Exomes (PAGES) group.[4] Genes were selected for this fetal anomalies panel if there was a high likelihood that the associated phenotype was likely to present during fetal development.
2) Intrauterine growth retardation gene set consisting of 611 genes mapped to the HPO term, 'intrauterine growth retardation', by the Human Phenotype Ontology project.[5] Intrauterine growth retardation is defined as an abnormal restriction of fetal growth with fetal weight below the tenth percentile for gestational age. For the intrauterine growth restriction and prematurity-associated gene set, the Human Phenotype Ontology project used text mining of Online Mendelian Inheritance in Man (OMIM)[6] to curate the gene lists.
3) Prematurity-associated gene list consisting of 170 genes mapped to the HPO term, 'premature birth', by the Human Phenotype Ontology project.[5]
4) A set of 221 genes previously associated with stillbirth compiled curated by Stanley *et al*.[7]

#### Association between gestational duration and non-transmitted alleles

To determine whether the association between gestational duration and selected PGS reflects direct genetic effects or the influence of non-transmitted parental alleles, we applied a "trio model"[8] to a subset of probands from DDD ($N_{trios}$=3,101) and 100kGP ($N_{trios}$=2,989). This model estimates the association of the phenotype with both transmitted alleles (child's PGS, $PGS_T$) and non-transmitted parental alleles ($PGS_{NT}$), allowing the direct genetic effect to be calculated as $\delta = \theta_T - \theta_{NT}$ in the following regression[8]:

$$Phenotype_{child} \sim \hat{\theta}_T \times PGS_T + \hat{\theta}_{NT} \times PGS_{NT}$$

This regression can be decomposed further to:

$$Phenotype_{child} \sim (\hat{\theta}_T - \hat{\theta}_{NT}) \times PGS_{child} + \hat{\theta}_{NT} \times (PGS_{mother} + PGS_{father})$$

In this model, the coefficient of the child's PGS represents the direct genetic effect, while the coefficient of the parents' PGS reflects the association of non-transmitted alleles with the outcome.

In our analysis, we estimated the direct genetic effect (δ) of the proband's PGS and the effects of the parental non-transmitted alleles ($\hat{\theta}_{m,NT}$ and $\hat{\theta}_{f,NT}$) on gestational duration using the following regression equation:

$$Gestational\ duration_i \sim \hat{\delta} \times PGS_i + \hat{\theta}_{m,NT} \times PGS_{i,m} + \hat{\theta}_{f,NT} \times PGS_{i,p} + sex_i$$

Here, $PGS_i$ represents the PGS for proband i, $PGS_{i,m}$ and $PGS_{i,p}$ are the PGS for proband's mother and father, respectively. The first 20 genetic principal components were regressed out of the PGS, and the residualised PGS were used in the analysis.

The model was initially run using data from all probands, followed by stratified analyses for diagnosed and undiagnosed probands. Analyses were performed independently for DDD and 100kGP, and effect size estimates from the two cohorts were meta-analysed using an inverse-variance weighted approach.

#### Deviations from pre-registered analysis plan

| Pre-registered method | Deviation | Justification |
|---|---|---|
| Testing 19 organ system-level HPO terms (present in >5% of probands) for associations with prematurity | Selected 13 organ system-level HPO terms for analysis | Excluded the HPO terms, “abnormality of the endocrine system”, “growth abnormality” and “neoplasm” as these terms were present in <5% of DDD probands. Used the HPO term “abnormality of the musculoskeletal system” instead of the descendant terms “abnormality of the skeletal system”, “abnormality of the musculature” and “abnormality of the connective tissue”. |
| Testing 14 medically relevant HPO-terms for an association with degree of prematurity | Excluded three of the proposed HPO terms from the analysis. | The model failed to converge for the HPO-terms “autistic behaviour”, “aggressive behaviour” and “hyperactivity” due to low numbers of probands with the HPO terms in certain gestational age groups. |

| | | |
|---|---|---|
| Definition of a monogenic diagnosis in GEL 100K. We will define "diagnosed" probands as those with a variant that is annotated as pathogenic or likely pathogenic and the family classified as being 'solved' or 'partially solved' by the referring clinician. All other probands that were included in the exit questionnaire will be classified as "undiagnosed". | We also classified probands who have a variant prioritised by the 100k Clinical Research Team, awaiting clinical review, in the 'Diagnostic Discovery data' as "diagnosed". | There is a high probability that the variants awaiting clinical review will be classified as disease causing.[9] |
| PGS will be calculated using summary statistics from the latest population-based GWAS for EA[10] ($PGS_{PopEA}$), NonCogEA[11] ($PGS_{NonCogEA}$), and CogEA[11] ($PGS_{CogEA}$). | Calculation of an additional PGS for gestational duration [12]. | Calculating a PGS for the phenotype of interest, gestational duration, facilitated a comparison between the effect of common variant predisposition to gestational duration and the effect of common variant predisposition to related traits, educational attainment and its cognitive and non-cognitive components, on gestational duration. |
| We will assess if the presence of a monogenic diagnosis modifies the association between PGS or DRVB and gestational duration by re-estimating the models and including an interaction between PGS, RVBS, and genetic diagnosis. | We tested for an association between the genetic measures in all probands, as well as in diagnosed and undiagnosed probands separately. | The revised analytical approach improved our ability to interpret the impact of a monogenic diagnosis on the relationship between selected genetic measures and gestational duration. This structure could also be applied to the trio models (described below) without requiring interaction terms between the genetic measure and each trio member's score for that measure, thereby simplifying the model. |
| We will test for an association between gestational duration, measured in weeks, and four PGS ($PGS_{PopEA}$, $PGS_{NonCogEA}$, $PGS_{CogEA}$) as well as DRVB by fitting linear regression models. | Additional polygenic score analysis was conducted to test for direct genetic effect of polygenic scores and DRVB on gestational duration in genotyped trios. | Associations between gestational duration and the proband's PGS for education related traits are likely to reflect associations between parental genetic predisposition to these traits and gestational duration. We expanded our analysis to a trio model to directly test whether the association between genetic measures and gestational duration was due to direct genetic effects and/or the effects of parental non- |

| | | |
|---|---|---|
| | | transmitted alleles. |
| We will use Fisher's exact tests with post-hoc pairwise comparisons to test for an enrichment of monogenic diagnoses in specific gene sets across the four gestational duration groups (term, moderately premature, very premature, and extremely premature) in DDD and GEL. | We combined the very or extremely preterm probands into one gestational age group for this analysis. | We opted to combine the very and extremely preterm probands into one group due to the small number of diagnosed probands in these gestational age groups. |
| No analysis of *de novo* variation | We tested for an enrichment of pathogenic DNMs exome-wide and in genes with and without a known association with DDs in preterm and term probands. We compared the attributable fraction between the gestational age groups and tested for genes enriched for DNMs in preterm and term probands. | This analysis allowed us to better understand the contribution of DNMs to DDs in preterm vs. term probands. Further, we were able to identify genes enriched for DNMs among preterm probands. |

#### Supplementary Figures

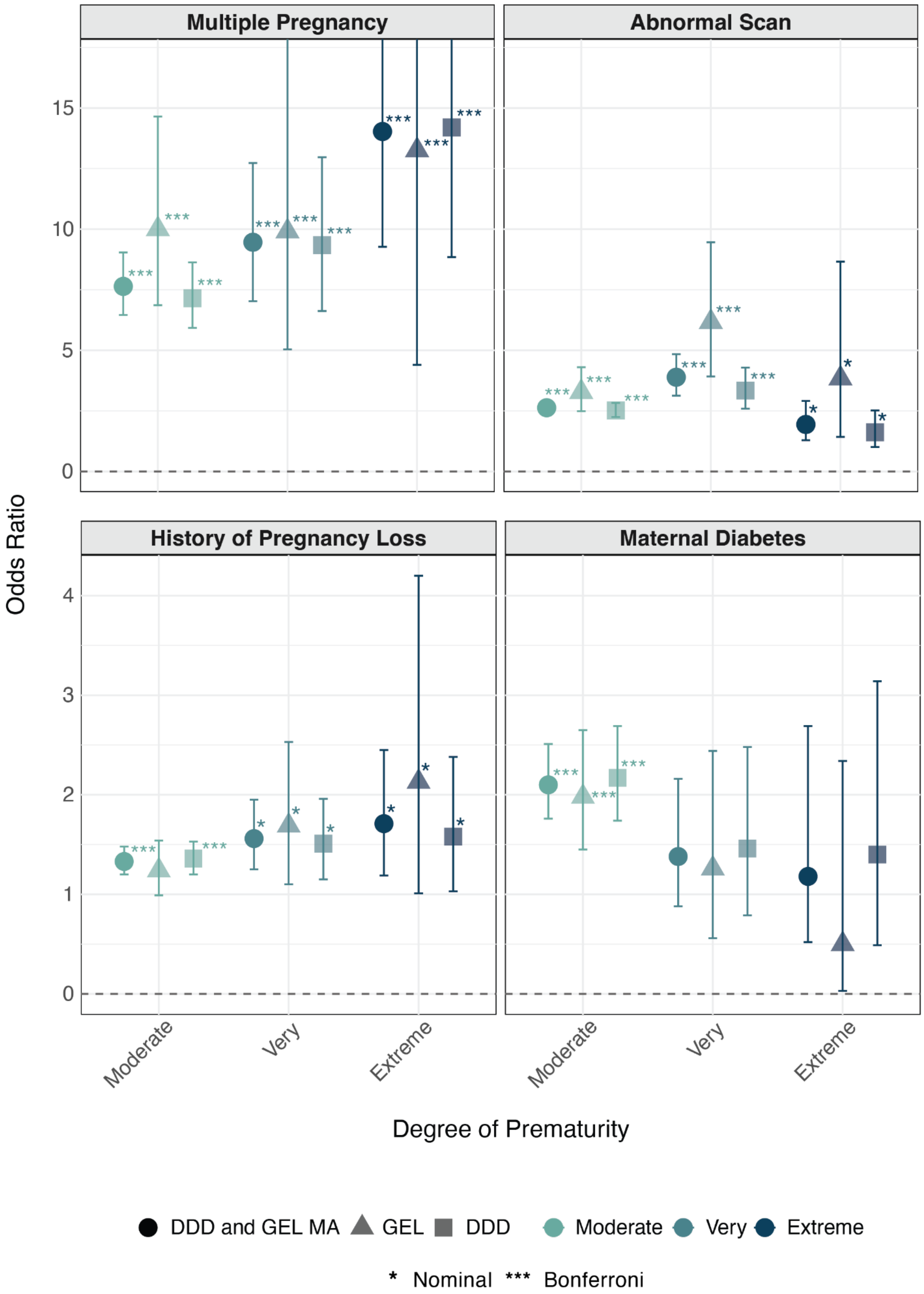


**Figure S1. Associations between antenatal factors and degree of prematurity.** The odds of a reported antenatal factor in preterm probands compared to term probands. Odds ratios are presented for each degree of prematurity relative to term probands, including results from the meta-analysis (MA) and cohort-specific analyses. Error bars indicate 95% confidence intervals.

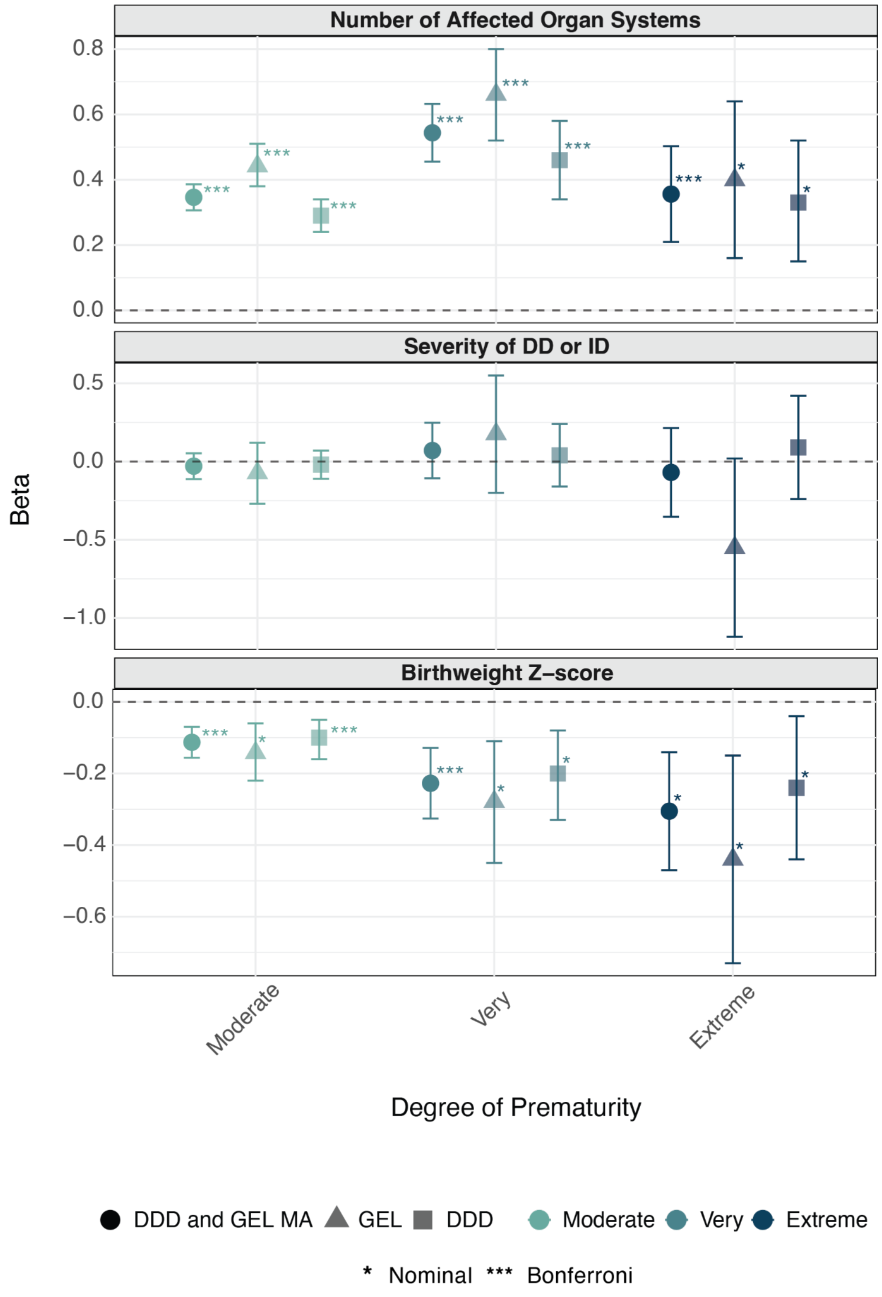


**Figure S2. Associations between selected clinical outcomes and degree of prematurity.** The figure depicts the standardised effect of each degree of prematurity on selected clinical outcomes compared to being born at term. The figure shows results from the meta-analysis (MA) and cohort-specific analyses. Error bars indicate 95% confidence intervals.

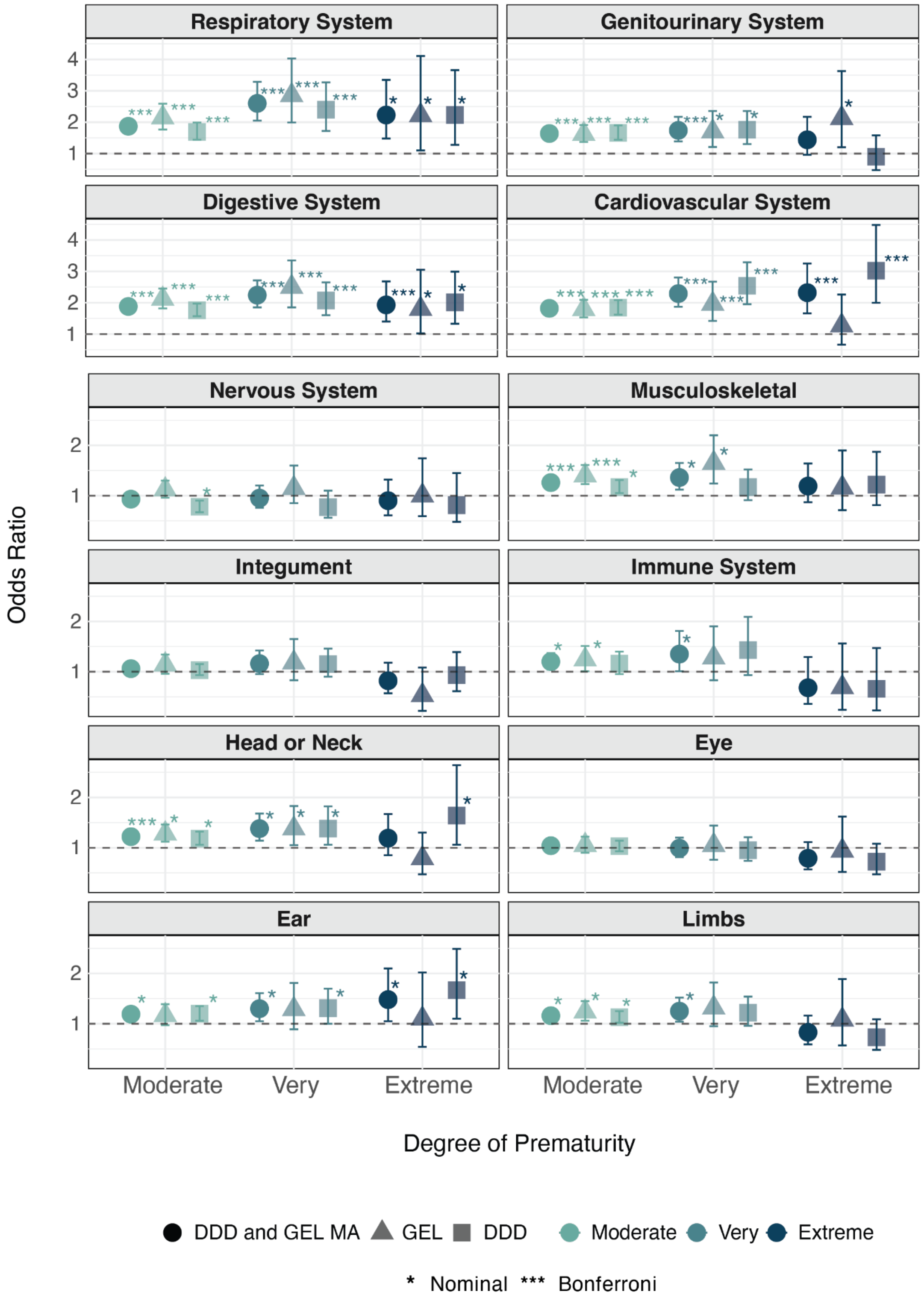


**Figure S3. Associations between abnormalities of an organ/organ-system and degree of prematurity.** The figure depicts the odds of being assigned an HPO term related to an abnormality of an organ or organ-system for each degree of prematurity compared to term. The figure shows results from the meta-analysis (MA) and cohort-specific analyses. Error bars indicate 95% confidence intervals.

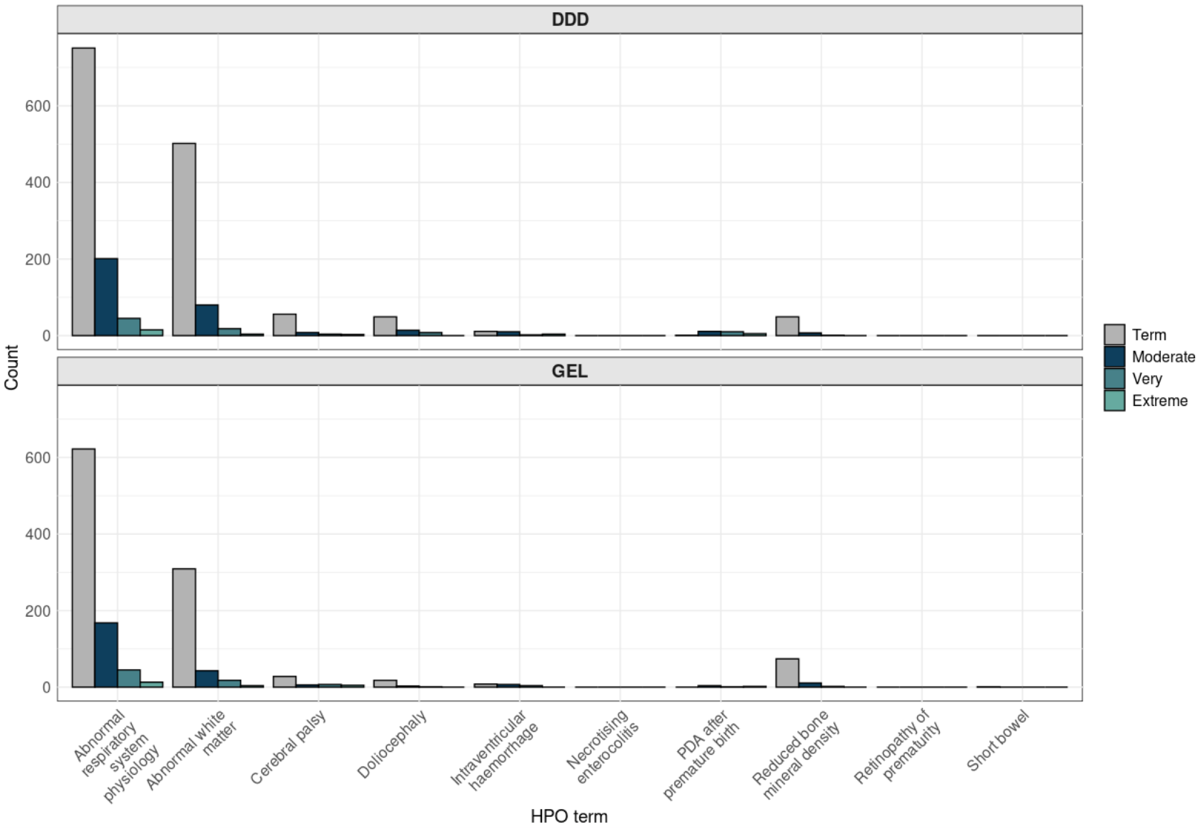


**Figure S4. Probands in DDD and GEL with prematurity-associated HPO terms.** Number of probands per a gestational age group with an HPO term from a list of terms curated by clinicians for a high likelihood of association with preterm birth.

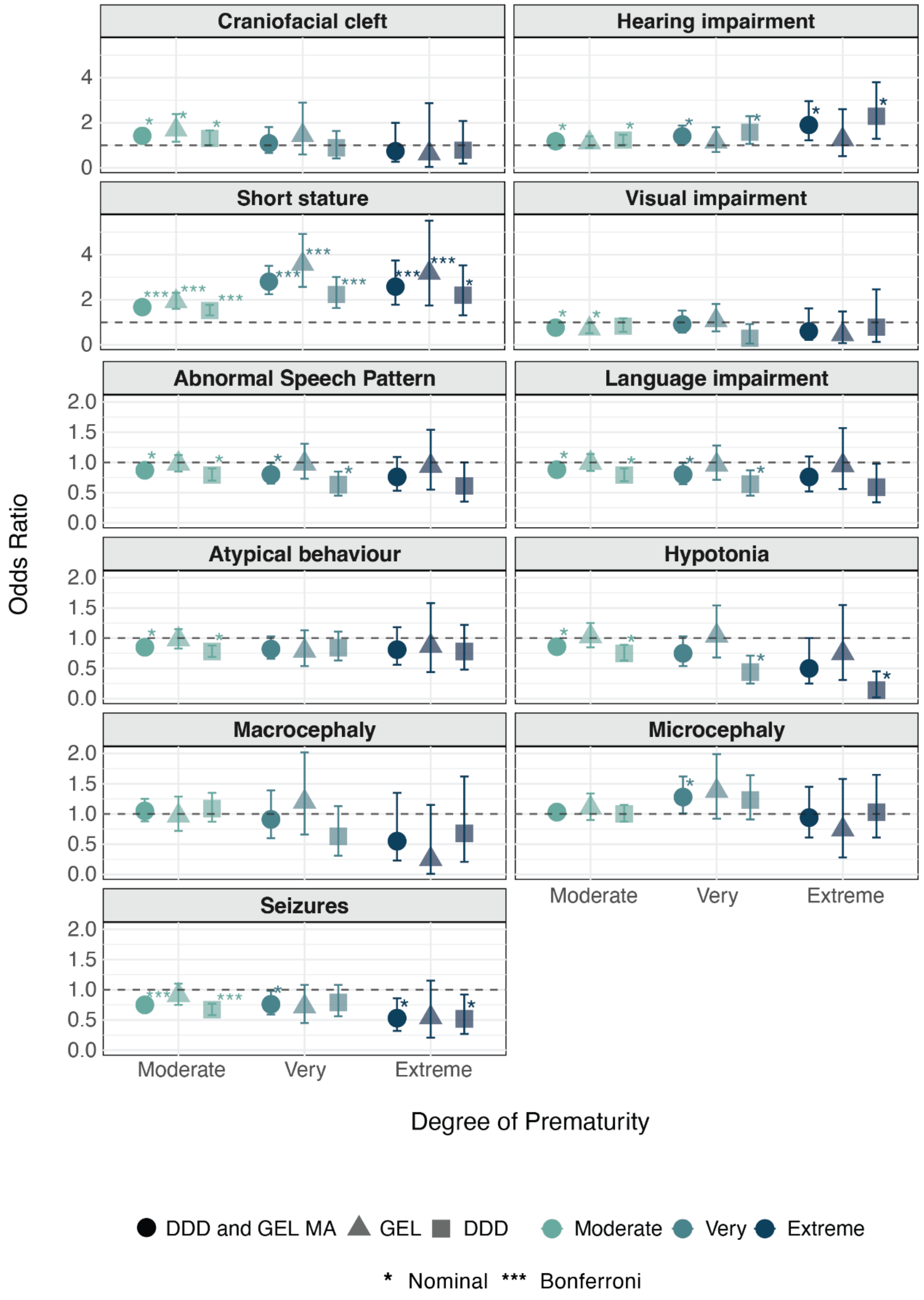


**Figure S5. Associations between medically-relevant HPO terms and degree of prematurity.** The figure depicts the odds of being assigned a medically-relevant HPO term for each degree of prematurity compared to term. The figure shows results from the meta-analysis (MA) and cohort-specific analyses. Error bars indicate 95% confidence intervals.

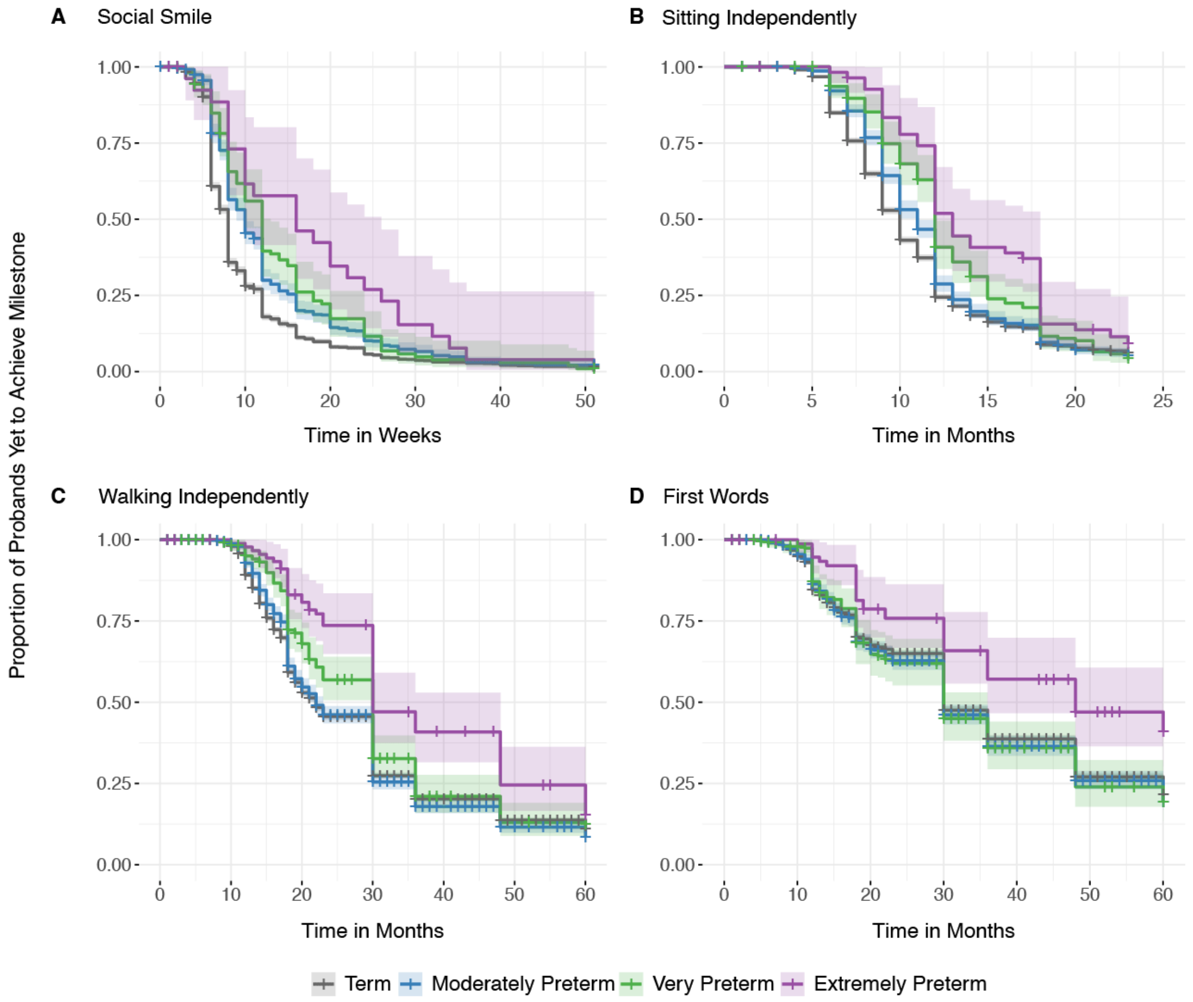


**Figure S6. Kaplan Meier curve comparing time taken to achieve developmental milestones across the gestational age groups.** X-axis denotes time in weeks or months, 0 corresponds to date of birth, and the y-axis shows the proportion of probands who have not achieved the developmental milestone after a given time period. **A)** Time (in weeks) taken to achieve the developmental milestone, social smile. **B**) Time (in months) taken to achieve the developmental milestone, sitting independently. **C**) Time (in months) taken to achieve the developmental milestone, walking independently. **D**) Time (in months) taken to achieve the developmental milestone, first words. Colour represents gestational age group; shaded ribbons depict 95% confidence intervals.

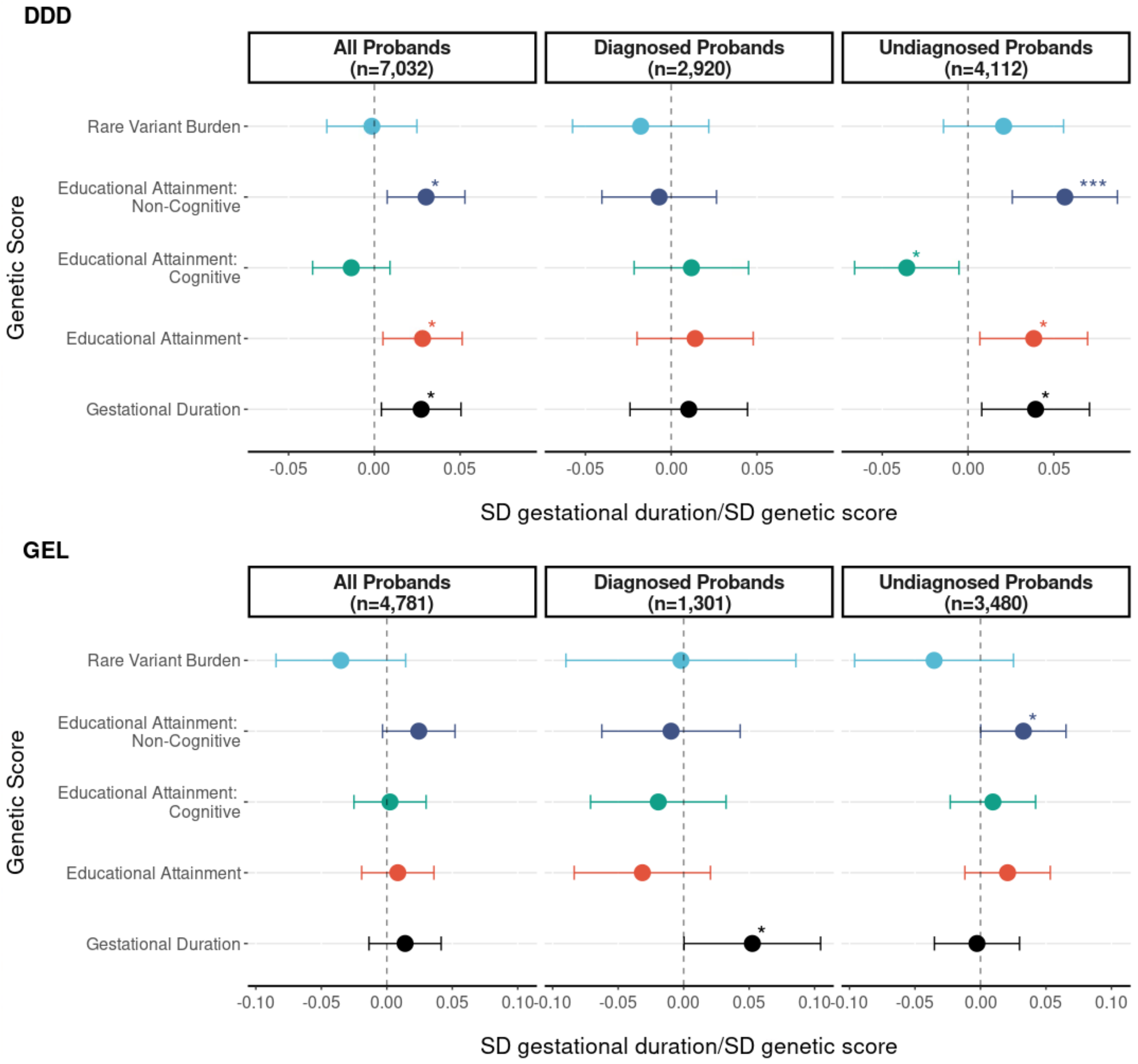


**Figure S7. Associations between proband PGS and gestational duration in DDD and GEL 100kGP.** Standardised effect of proband's genetic measure on gestational duration. Gestational duration was rank-based inverse normal transformed (RINT), the beta should be interpreted as change in standard deviation of RINT gestational duration per standard deviation change in genetic score. Top panel shows estimates for DDD probands and bottom panel for 100kGP. Error bars represent 95% confidence intervals. Three asterisks indicate that the association passed Bonferroni correction for multiple testing, one asterisk indicates nominal significance ($p<0.05$).

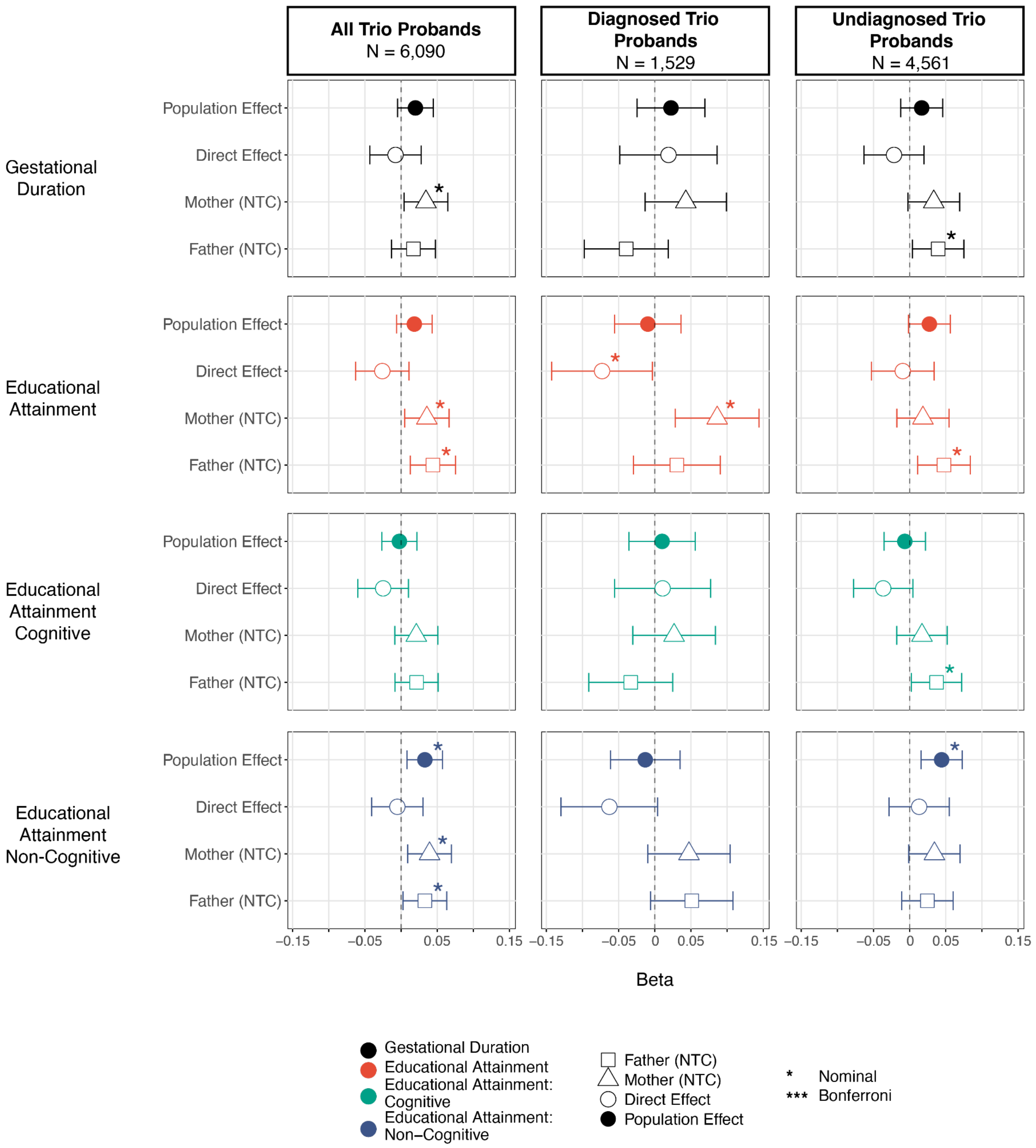


**Figure S8. Associations Between PGS and Gestational Duration in meta-analysis of DDD and GEL 100kGP Trios.** Effect size estimates are shown before controlling for parental PGS ("population effect") and after controlling for parental PGS ("direct effect") in full trios with genetic data. Gestational duration was rank-based inverse normal transformed (RINT), the beta should be interpreted as change in standard deviation of RINT gestational duration per standard deviation change in PGS. Error bars represent 95% confidence intervals.

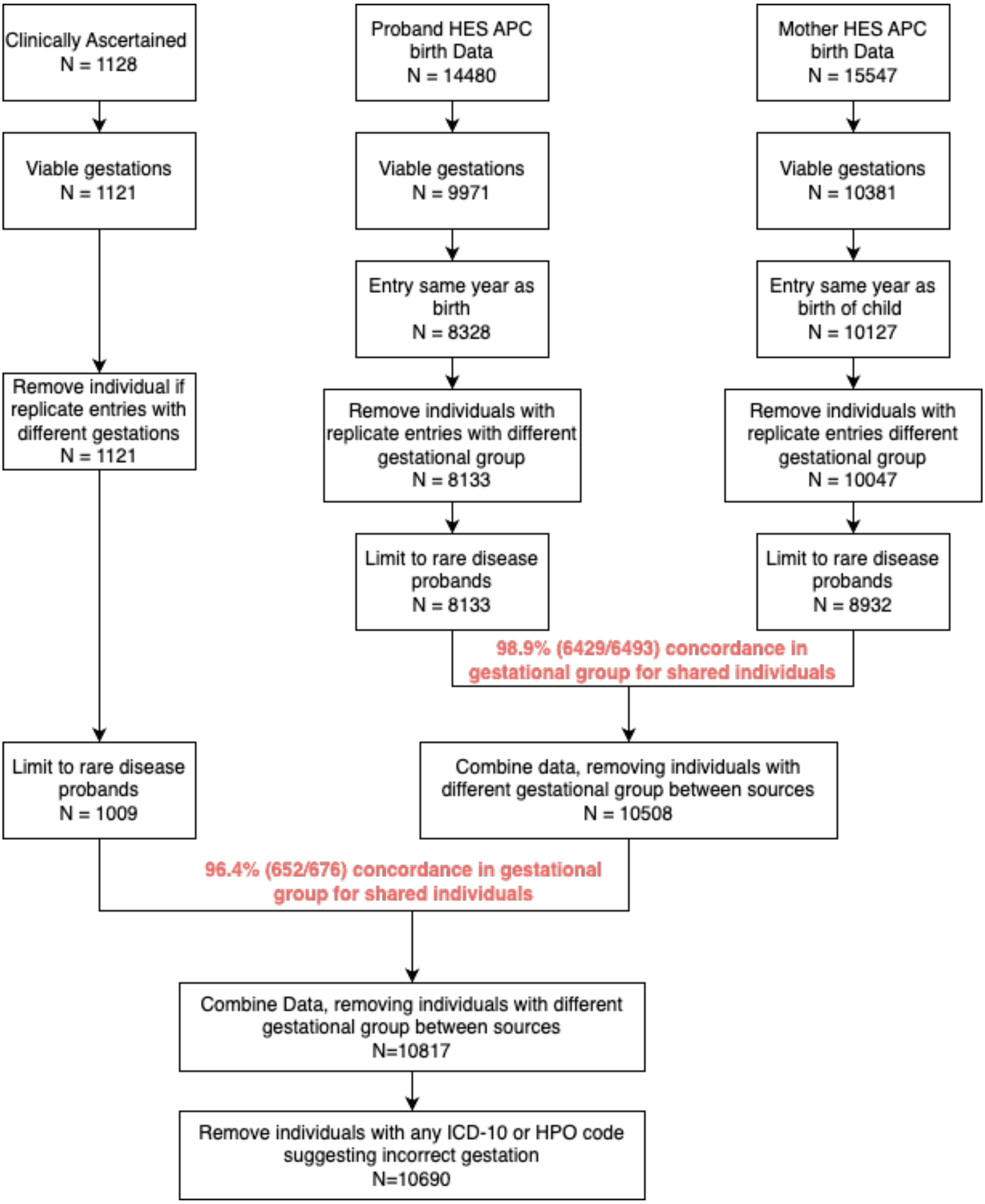


**Figure S9. Gestational duration data filtering flowchart in the 100kGP for all rare disease probands.**

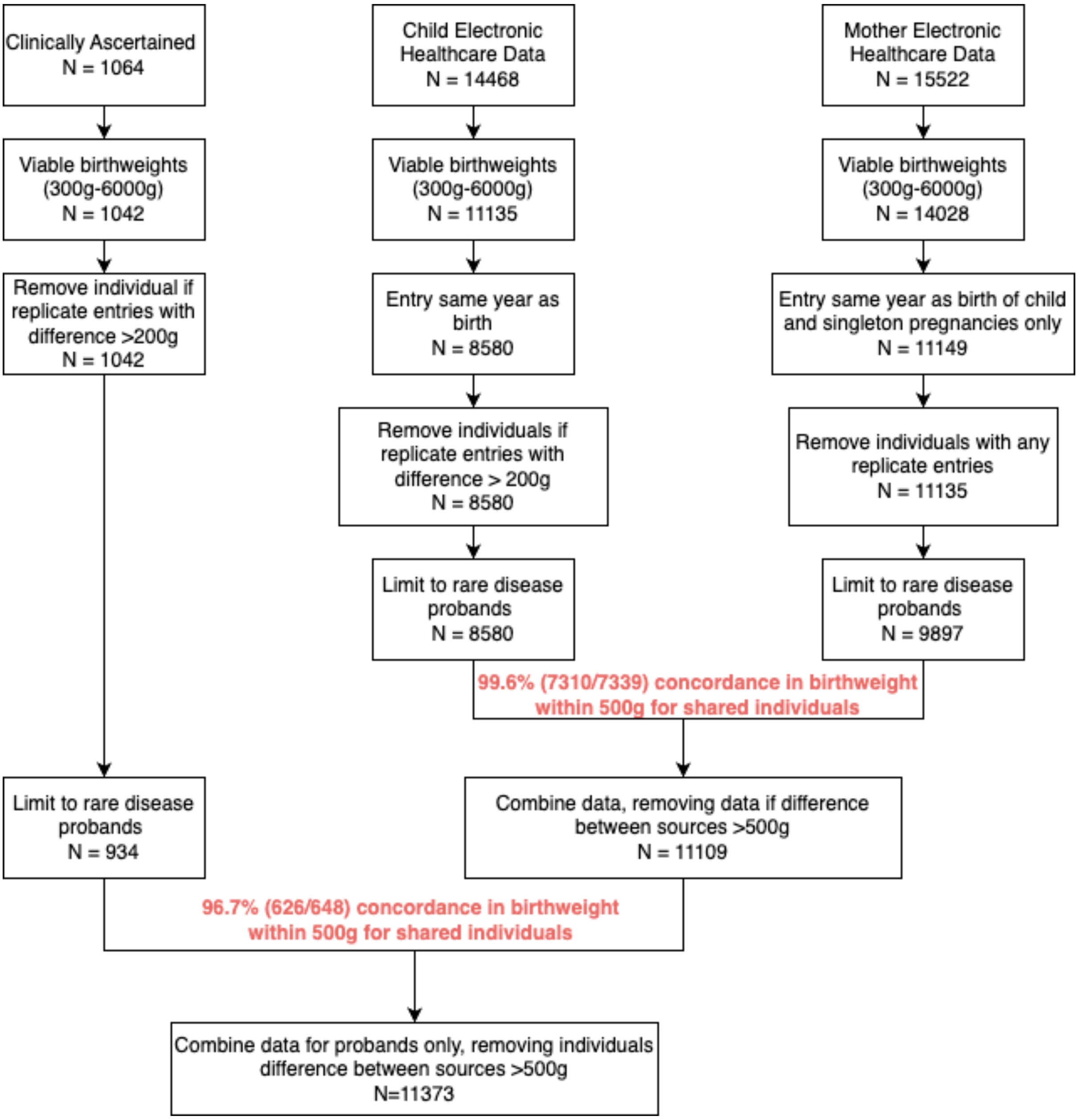


**Figure S10. Birthweight data filtering flowchart in the 100kGP for all rare disease probands.**

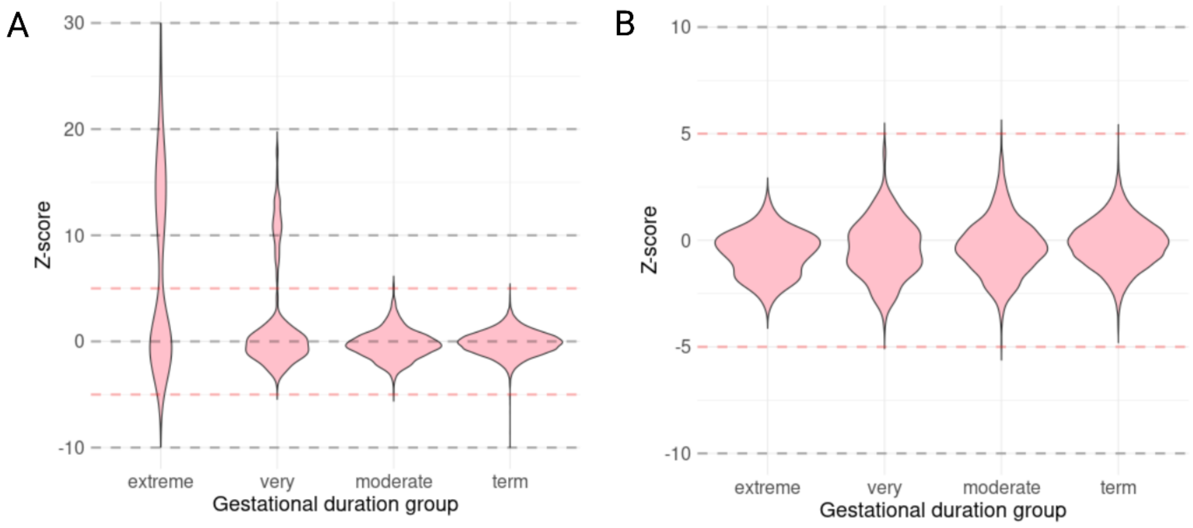


**Figure S11. Z-score of birthweights in the 100kGP.** A) Z-scores based on gestational duration and sex for all rare disease probands. B) Z-scores for all rare disease probands after removing individuals with z-scores greater than 5.

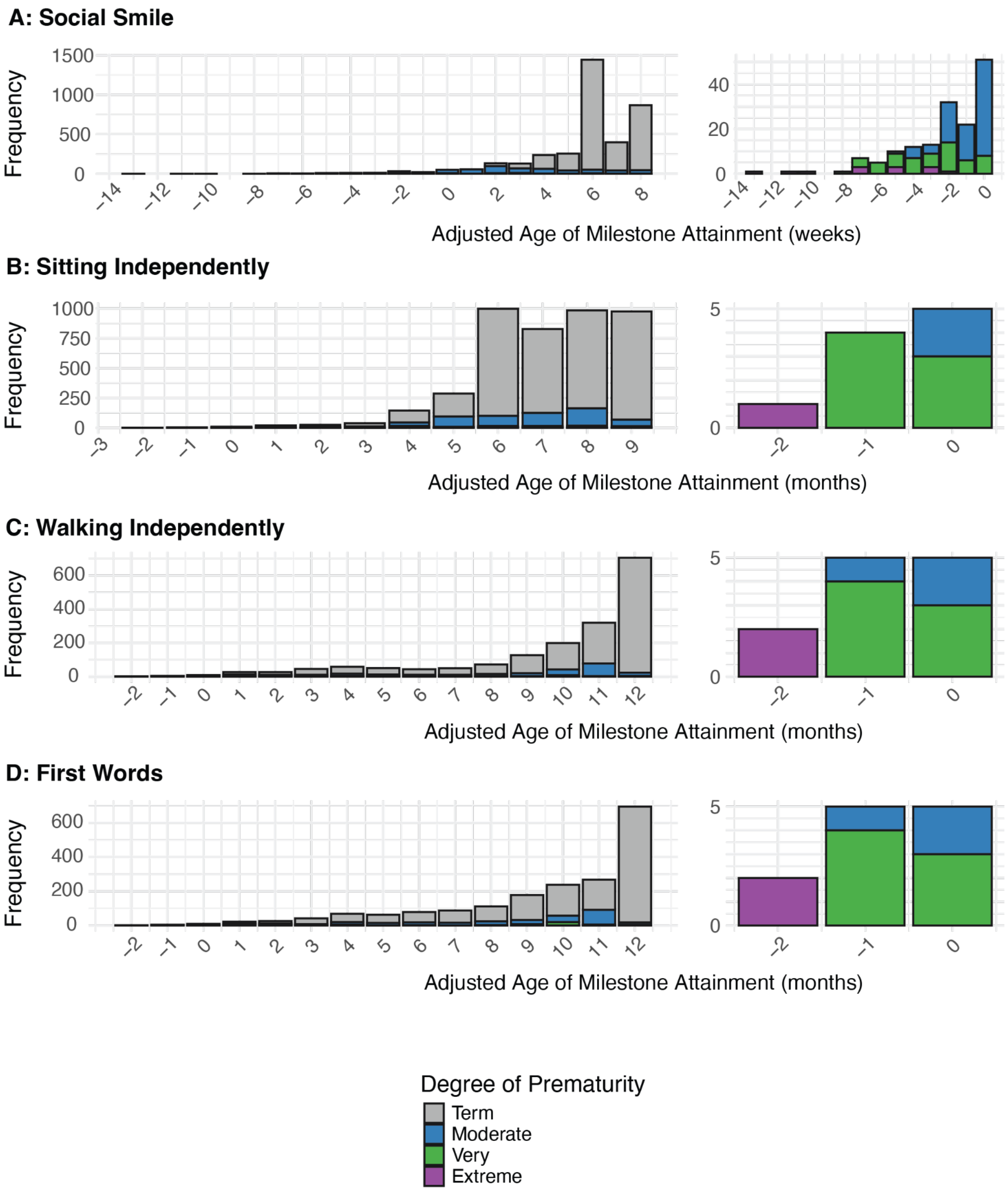


**Figure S12. Stacked bar chart showing corrected age of developmental milestone attainment per gestational age group.** The figure shows the number of probands in each gestational age group who attained a developmental milestone at a specific age (measured in weeks or months) after correction for gestational age at birth. Panels A–D present stacked bar charts for each assessed developmental milestone, with the panel on the right displaying the number of probands whose corrected milestone age indicates achievement prior to birth (i.e., at or before time point 0).

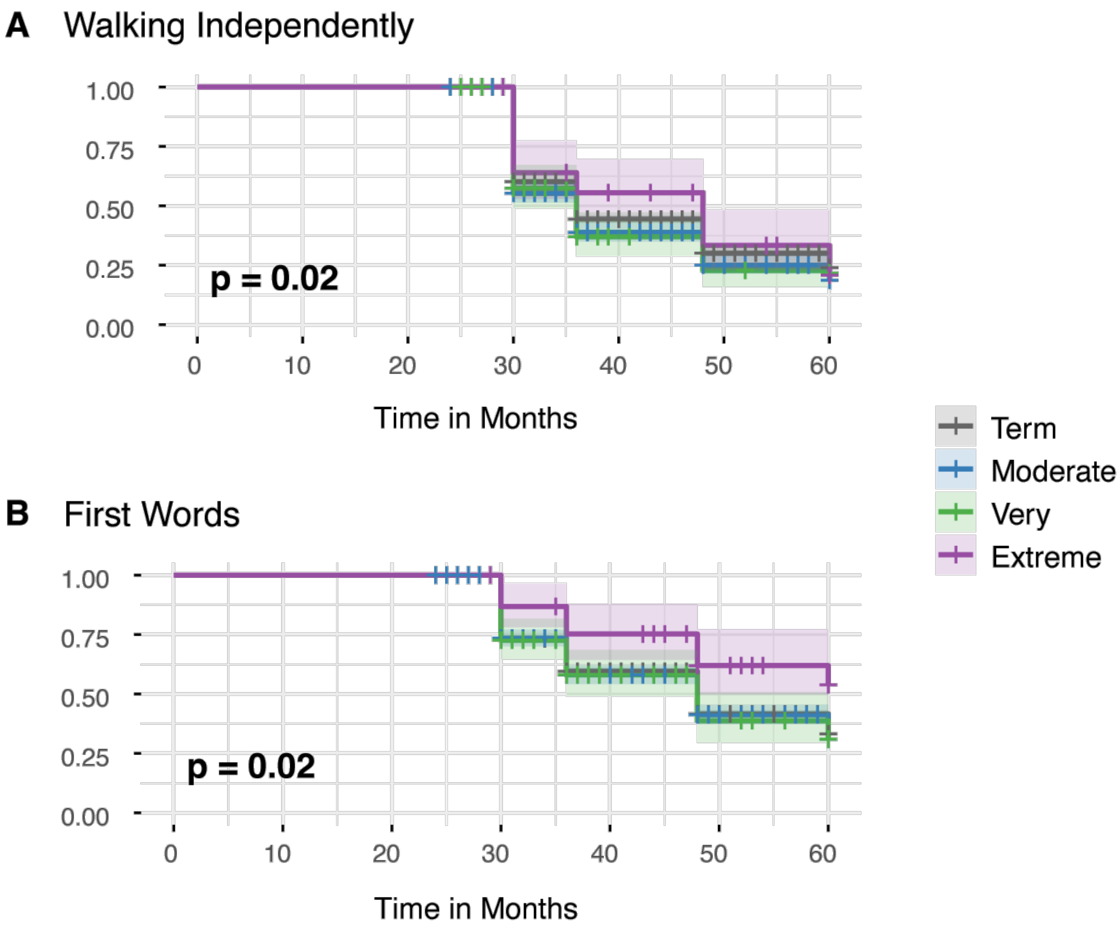


**Figure S13. Kaplan Meier curve comparing time taken to achieve developmental milestones across the gestational age groups for probands who attained the milestone after two years of age.** X-axis denotes time in months, 0 corresponds to date of birth, and the y-axis shows the proportion of probands who have not achieved the developmental milestone after a given time period. **A**) Time taken to achieve the developmental milestone, walking independently. **B**) Time taken to achieve the developmental milestone, first words. Colour represents gestational age group; shaded ribbons depict 95% confidence intervals. P-values from the log-rank test comparing survival curves across gestational age groups are provided for each milestone.

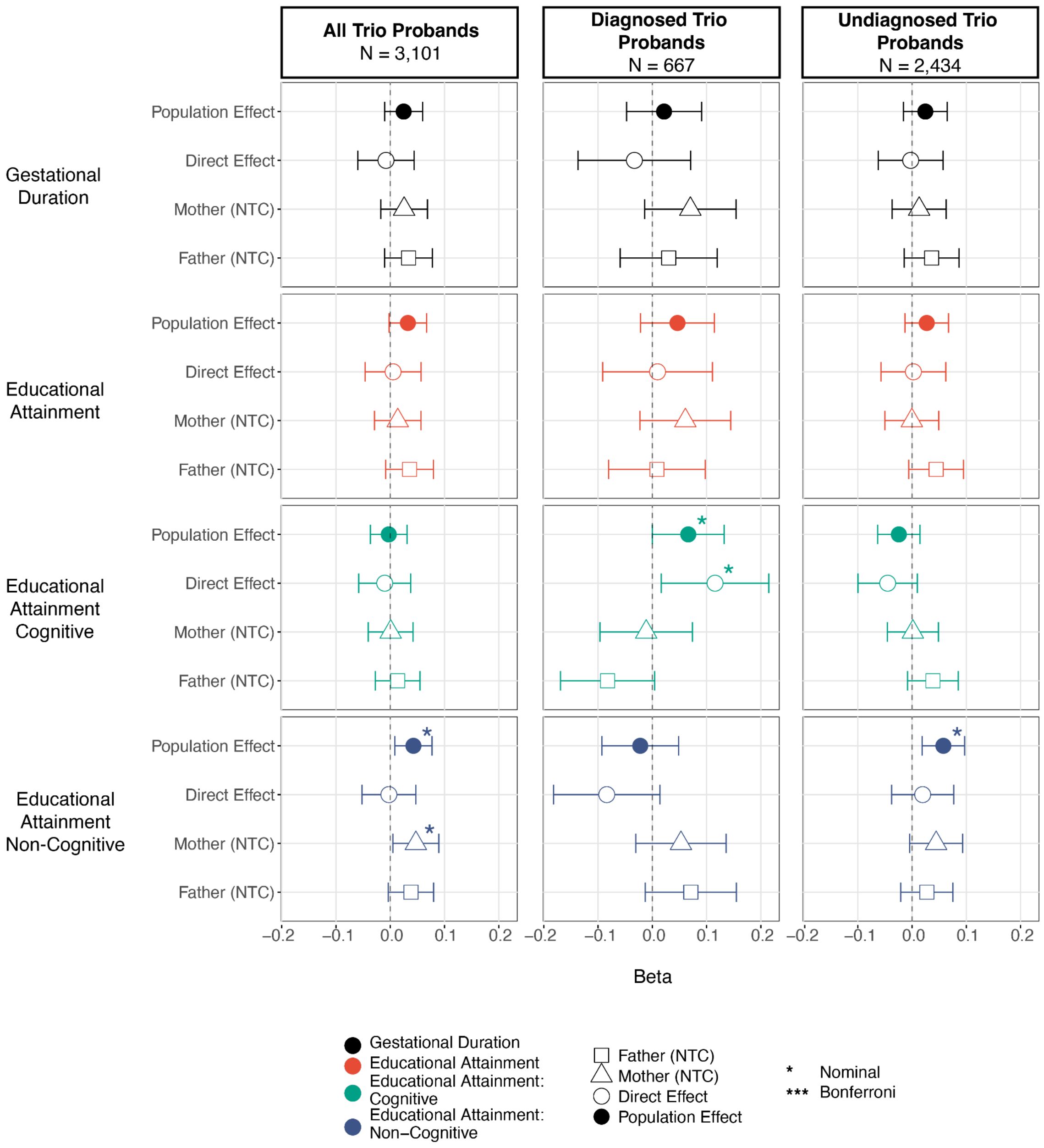


**Figure S14. Associations Between PGS and Gestational Duration in DDD Trios.** Effect size estimates are shown before controlling for parental PGS ("population effect") and after controlling for parental PGS ("direct effect") in full trios with genetic data. Gestational duration was rank-based inverse normal transformed (RINT), the beta should be interpreted as change in standard deviation of RINT gestational duration per standard deviation change in PGS. Error bars represent 95% confidence intervals.

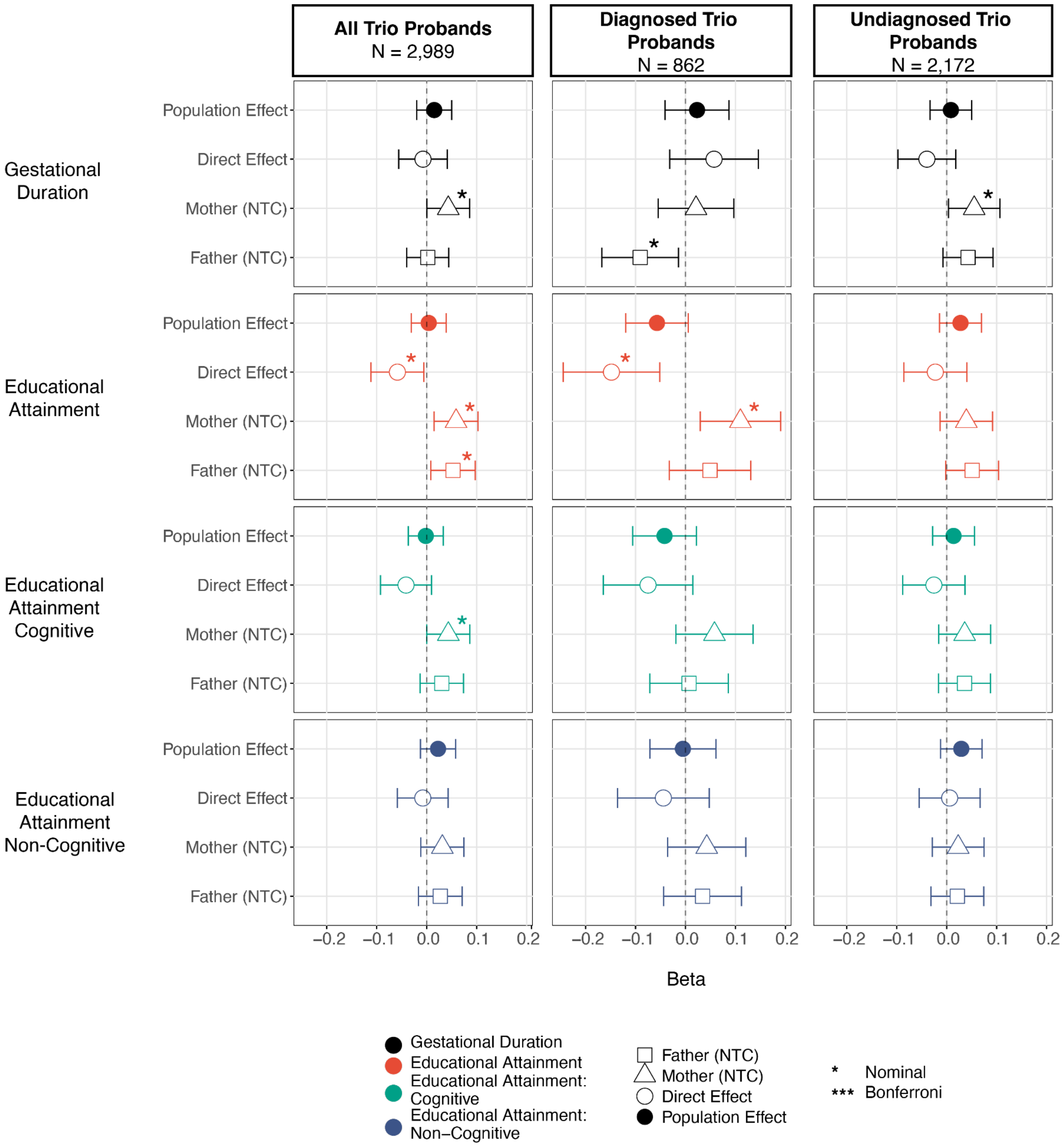


**Figure S15. Associations Between PGS and Gestational Duration in GEL 100kGP Trios.** Effect size estimates are shown before controlling for parental PGS ("population effect") and after controlling for parental PGS ("direct effect") in full trios with genetic data. Gestational duration was rank-based inverse normal transformed (RINT), the beta should be interpreted as change in standard deviation of RINT gestational duration per standard deviation change in PGS. Error bars represent 95% confidence intervals.

### Supplementary Notes

#### Supplementary Note 1: Quality control of DDD developmental milestone data

As part of the assessment of DDD probands, recruiting clinicians recorded the age at which each proband achieved developmental milestones in DECIPHER. In clinical practice, the reported age of milestone attainment is adjusted for gestational age until two years of age in children born preterm.[13] In some cases, it was speculated that clinicians may have already made such adjustments for premature probands even though this was not specifically requested. To explore this, we conducted an analysis of the developmental milestone data. First, we corrected the developmental milestones for gestational age at birth by subtracting the number of weeks born preterm from the age of reported milestone attainment in all premature probands. After performing this correction, we found that the "corrected" age of milestone attainment was less than zero for several probands, implying that the milestone was achieved before birth (**Supplementary Figure 12**). This finding suggests that, for at least some premature probands, the recruiting clinician recorded the corrected age of milestone attainment. For this reason, we chose not to correct the milestone data for gestational age in our main analyses. However, as a sensitivity analysis, we repeated the Kaplan-Meier survival analysis restricting to probands who achieved the milestones of walking independently and first words after two years of age, the age beyond which correction is no longer performed. We did not repeat the analysis for social smile and sitting independently, as no probands were reported to achieve these milestones after two years of age. The log rank test from the Kaplan-Meier analysis showed a significant difference in the time taken to achieve the two developmental milestones across gestational age groups in DDD (log rank test $p=2x10^{-2}$)(**Supplementary Figure 13**). As in the analysis including all probands, the difference across gestational age groups appears to be primarily driven by delays among the extremely preterm probands. The findings from the repeat survival analysis align with those of the initial analysis. Together, these results support the conclusion that prematurity, particularly extreme prematurity, delays development among probands with DDs suspected to have a genetic cause.

#### Supplementary Note 2: Association between rare variant burden score and gestational duration

Rare damaging coding variants are associated with NDD risk as well as lower educational attainment in the general population.[14–16] We thus hypothesised that rare genetic variation may also affect associations between gestational duration and phenotypic presentation in DDs. To explore this, we tested for an association between the number of inherited damaging coding variants in loss-of-function-intolerant genes, captured as a rare variant burden score (RVBS), and gestational duration. We excluded rare *de novo* variants from the RVBS as these variants are more likely to be fully diagnostic, since most parents in this cohort are clinically unaffected, whereas rare inherited variants are likely to be associated with reduced educational attainment in the parents. We found no associations between RVBS and gestational duration in the meta-analysis or in the cohort-specific analyses. Therefore, there is currently no significant evidence that deleterious rare variants inherited from parents influence gestational duration. However, it seems likely that future better-powered studies in population-based cohorts will allow the discovery of rare variants affecting gestational timing.

#### Supplementary Note 3: Association between non-transmitted alleles and gestational duration

We hypothesised that the observed associations between the proband's PGS for EA, the non-cognitive component of EA, and gestational duration were not due to direct genetic effects. Instead, these associations might be driven by other factors correlated with the child's genotype, such as parental alleles that influence the phenotype through the shared environment, including alleles that are not transmitted to the child. To explore this, we fit a trio model, in which the phenotype (gestational duration) is regressed on both the child's PGS and the parental PGSs, in a subset of probands with genetic data for both parents. In this model, the coefficient on the child's PGS represents the direct genetic effects, whereas the coefficients of the parental PGS capture the association between gestational duration and the parental alleles that are not transmitted to the offspring and are thus referred to as "non-transmitted coefficients". We found no evidence for a direct genetic effect of any of the PGS on gestational duration in the meta-analysis of all probands (**Supplementary Figure 8**). Rather, we observed a nominally significant ($p < 0.05$) positive association between gestational duration and the parental non-transmitted alleles for EA and its non-cognitive component, as well as with the maternal non-transmitted alleles for gestational duration (**Supplementary Figure 8**). When stratifying by the proband's diagnostic status, many of the observed associations between gestational duration and the non-transmitted coefficients became null, possibly due to limited power for estimating the trio model in these subsets. Similarly, in the cohort-specific analysis, no associations were consistently observed across cohorts, which may reflect both limited power and differences in trio ascertainment (**Supplementary Figure 14**, **Supplementary Figure 15**).
